# Pneumask: Modified Full-Face Snorkel Masks as Reusable Personal Protective Equipment for Hospital Personnel

**DOI:** 10.1101/2020.04.24.20078907

**Authors:** Laurel Kroo, Anesta Kothari, Melanie Hannebelle, George Herring, Thibaut Pollina, Ray Chang, Dominic Peralta, Samhita P. Banavar, Eliott Flaum, Hazel Soto-Montoya, Hongquan Li, Kyle Combes, Emma Pan, Khang Vu, Kelly Yen, James Dale, Patrick Kolbay, Simon Ellgas, Rebecca Konte, Rozhin Hajian, Grace Zhong, Noah Jacobs, Amit Jain, Filip Kober, Gerry Ayala, Quentin Allinne, Nicholas Cucinelli, Dave Kasper, Luca Borroni, Patrick Gerber, Ross Venook, Peter Baek, Nitin Arora, Philip Wagner, Roberto Miki, Jocelyne Kohn, David Kohn Bitran, John Pearson, Cristián Muñiz Herrera, Manu Prakash

## Abstract

Here we adapt and evaluate a full-face snorkel mask for use as personal protective equipment (PPE) for health care workers, who lack appropriate alternatives during the COVID-19 crisis in the spring of 2020. The design (referred to as Pneumask) consists of a custom snorkel-specific adapter that couples the snorkel-port of the mask to a rated filter (either a medical-grade ventilator inline filter or an industrial filter). This design has been tested for the sealing capability of the mask, filter performance, CO2 buildup and clinical usability. These tests found the Pneumask capable of forming a seal that exceeds the standards required for half-face respirators or N95 respirators. Filter testing indicates a range of options with varying performance depending on the quality of filter selected, but with typical filter performance exceeding or comparable to the N95 standard. CO2 buildup was found to be roughly equivalent to levels found in half-face elastomeric respirators in literature. Clinical usability tests indicate sufficient visibility and, while speaking is somewhat muffled, this can be addressed via amplification (Bluetooth voice relay to cell phone speakers through an app) in noisy environments. We present guidance on the assembly, usage (donning and doffing) and decontamination protocols. The benefit of the Pneumask as PPE is that it is reusable for longer periods than typical disposable N95 respirators, as the snorkel mask can withstand rigorous decontamination protocols (that are standard to regular elastomeric respirators). **With the dire worldwide shortage of PPE for medical personnel, our conclusions on the performance and efficacy of Pneumask as an N95-alternative technology are cautiously optimistic**.

**Key points:** Full-face snorkel masks are adapted for use as Personal Protective Equipment during the COVID-19 crisis, using a custom adapter that facilitates the attachment of inline medical-grade respiratory filters or NIOSH industrial respirator filters. This solution was designed as a reusable stopgap solution for healthcare workers to help address the short-term global N95 respirator shortage.

## 1 Description of Concept

Personal protective equipment (PPE) is one of the most important protective layers for healthcare workers around the world in a crisis like COVID-19 [1, 2]. However, the supply of PPE in hospitals is extremely limited and the crisis is worsening by the day [2-4], as the industrial supply chains are unable to scale up to meet current demand [5]. We propose a potential stop-gap solution which consists of three parts: an off-the-shelf snorkel mask, a custom (3D-printed / injection-molded) adapter, and a filter/filter cartridge. The primary benefit of a snorkel mask is providing a full-face shield and air seal while allowing for controlled intake and exhaust flows through rated respiratory filters. Such masks are already widely available in large quantities and designing to make use of this supply chain could allow better access to PPE for medical personnel during this crisis. Typical full-face snorkel masks have a respiratory snorkel-port located at the top of the mask consisting of 1 inhale and 2 exhale channels (Figure 1). In most models, there is an additional one-way exhale port near the mouth/chin area. The full-face mask has been tested to withstand disinfection protocols, while still maintaining its seal performance after the disinfection. This allows the mask to be safely reused, if proper standard operating procedures are followed.

**Figure 1.**
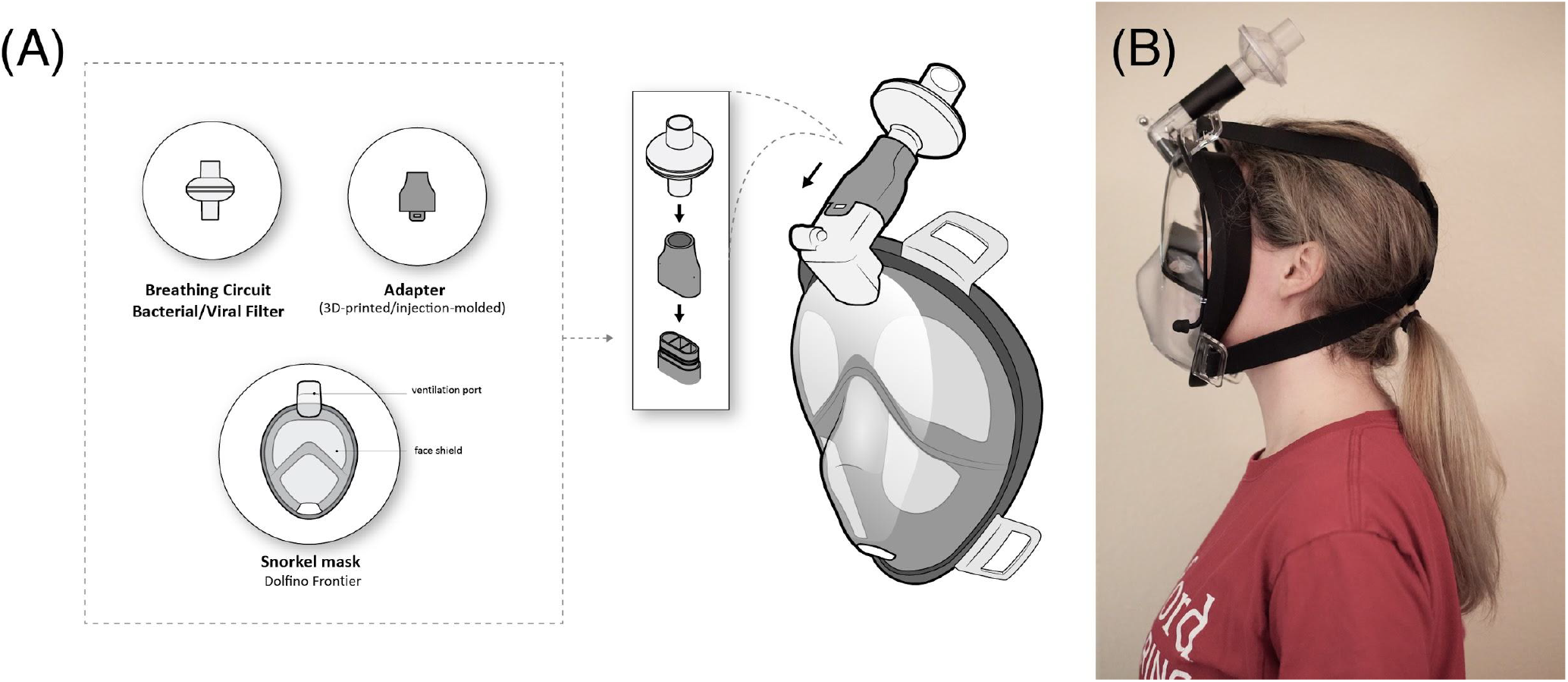
(A) Proposed Pneumask and (B) Prototype pictured on user. The snorkel mask is connected to an inline viral filter to provide protections to healthcare workers. The snorkel mask and adapter can be disinfected thus provide a reusable solution.

Our designs are aimed specifically at doctors, nurses and hospital staff (for use in settings where no alternative FDA-approved medical PPE is available or suitable for use). Unlike many other open-source designs that are intended for ventilated patients or consumers, this mask is primarily designed to prioritize the protection of a healthy user, rather than the surrounding environment. It is assumed that users are alert and active during mask usage, can be trained on standard operating procedures, and have access to standard sterilization and disinfection methods used in hospitals. Additionally, the appropriate filters for Pneumask are currently only accessible to those with a hospital affiliation (in the USA, at the time of this publication). This concept is not intended for patient use.

The goal is to couple the snorkel-port of the mask to a filter through a custom adapter. There are a couple of strategies with regards to the filter: (1) Rated off-the-shelf medical respiratory inline filters can be directly connected to the adapter, or (2) Alternative filter materials, such as NIOSH-rated industrial filters, can be attached with a second modular adapter. (A list of medical inline filters and industrial-used filters are listed in Appendix A Table 1 and Appendix A Table 4.) Many of these filters are rated for a substantially longer life-span as compared to the standard disposable N95 respirators.

This project is an effort to provide a safer solution to the current PPE shortage crisis compared to untested homemade masks or bandanas, which often cannot provide good seals around the face and are not tested for high filtration efficiency.

## 2 Design

### 2.1 Diversity of Full-Face Snorkel Masks

Today a huge diversity of full-face snorkel masks are commercially available (Please refer to Appendix A Table 3 to see the list of commercially available full-face snorkel masks). The original concept of full-face snorkel masks for recreational use came from the Subea Team (Figure S1). Subea has provided us a CAD file of their mask which enables a precise manufacture of an adapter. Additionally, we have also developed (reverse-engineered) the geometry of the connecting feature and the CAD adapter for the Dolfino Frontier mask, which is openly available. We have focused our design effort in this document specifically on the Dolfino Frontier mask unless otherwise stated. This choice to focus on the Dolfino Frontier mask was due to local availability in the USA at the start of this project.

### 2.2 Inhale-Exhale Pathways for the Snorkel Mask

To facilitate further design and communications, we have developed a standard nomenclature for possible connection configurations of a snorkel mask (Figure S6 in Appendix E). The original airflow pathway of a non-modified snorkel mask is shown in (Figure 2A,B), while the modifications to the flow path are depicted in 2B. In the normal mode of operation of a non-modified snorkel mask, the inhaled air passes through the center channel of the snorkel, enters the eyes chamber, then the mouth chamber, and is inhaled by the user. In water, the chin valve is blocked, and all the exhaled air goes through the two side channels of the snorkel at the top of the mask. In air, most of the exhaled air goes through the chin valve near the mouth.

**Figure 2.**
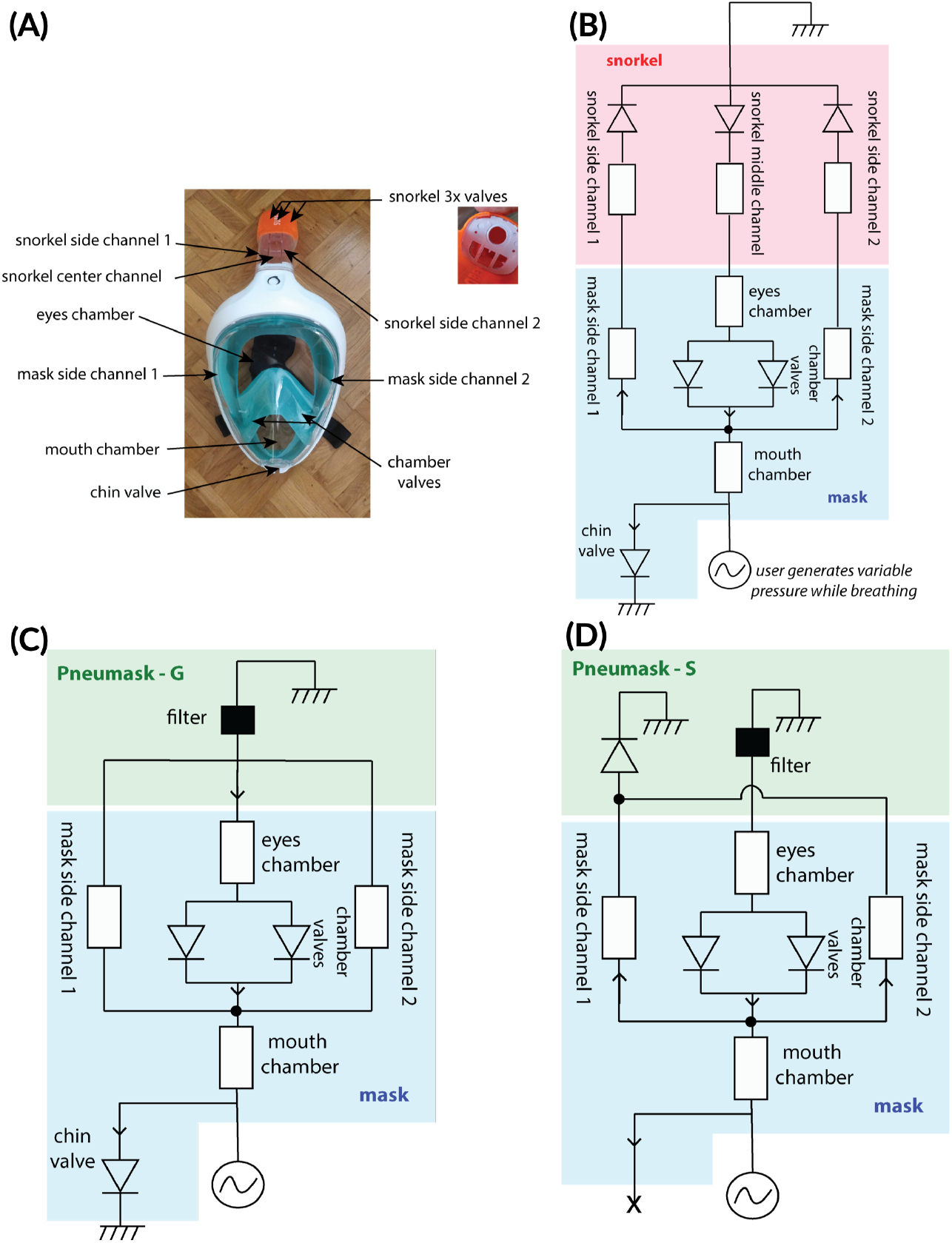
(A,B) Air flow pathways of the snorkel mask, based on Decathlon Easybreath, (C) Pneumask-G and (D) Pneumask-S. Notation: diodes=valves; ground=atmospheric pressure.

The modified flow path for use as PPE (general hospital usage), is very similar to the stock configuration of the mask: all inhale breath is directed through the filter, and the majority of exhale air is directed through the chin valve. We discuss alternative flow pathway designs specifically for surgeons.

### 2.3 Adapter Part Design

With aids of the airflow diagrams (See panels C and D of Figure 2), our team currently has two designs of the coupling adapters that are targeted at two different use-cases.

- ANESTHESIOLOGIST AND GENERAL-HOSPITAL USE PPE: The first, most prevalent use-case is a PPE solution for hospital personnel that do not require a “sterile-field”. Functionally, this means that the air exhaled by the user does not need to be filtered, or redirected away from the patient. This use-case is the application for the majority of the workers in hospitals. We will refer to this prototype in this document as “Pneumask-G”. (Figure 2C).
- SURGEON PPE: The second design is specifically targeting the needs of surgeons (a vast minority of the use-cases), which has filters on both the inhale and the exhale, and directs the exhale away from the patient. We will refer to this prototype in this document as “Pneumask-S”. (Figure 2D)

### 2.4 Pneumask-G: Anesthesiologist and General-Use Design

In Pneumask-G, the system is designed such that all inhaled air passes through the filter(s) attached to the top of the mask, and exhale is mostly channeled out through the built-in one-way valve at the chin of the mask (Figure 2C) with some exhale flowing out through the filter. The design of this main system is modular to allow adaptation to supply chain shortages of filters. It consists of 2 custom adapters:

1. The first piece is an adapter that couples the Dolfino Frontier full-face snorkel to a female filter port, specifically to accept ISO standard 22mm OD filters (Figure 3B). This adapter connects the snorkel to a single standard respiratory filter that is already in supply at many hospitals. All air from inhale is meant to be channeled through this adapter. An optional one-way valve can be placed between the adapter and the filter to prevent any exhaled air from going back to the filter, which lowers fogging and CO2 buildup. Optional valve and air pathway modifications are discussed in the adapter design supplementary. The final injection-molded adapter can be downloaded here (final version by Eric Gagner, .STEP only): https://www.dropbox.com/sh/sztddvy1or1auc7/AABI99D3pklqkQBGL7xdTKHNa?dl=0. The original 3D-printed design from the Prakash Lab is depicted in Figure 3c. Pull tests on this adapter were performed on several of our 3D-printed prototype models to ensure usability and secure fit. Our team tested the linear pull force necessary to disconnect the adapter/O-ring male mask coupling on a snorkel mask. We found this force to be 17lbs, which was the same as for the supplied snorkel, and compared favorably with our measured linear pull force necessary to disconnect the 22mm male ISO adapter/female ISO Virex filter coupling of 9lbs. The configuration of the Pneumask-G design shown in figure 3A can be implemented with any rated filter with an ISO 22mm OD port. This includes both HME filters (some of which are HEPA-rated) and viral filters. The performance of the design in terms of 1) work of breath and 2) CO2 accumulation will depend on the filter that is used inline in the system. Performance of different filters is discussed in the filter testing section. We found that it is possible to remove the soft side-channel tubes in the Dolfino Frontier mask, and “plug” the two remaining gaps that are left between the eye and mouth chambers. This can improve comfort slightly by forcing all exhaled air exclusively though the chin valve and decreasing inhale resistance, while also reducing backflow. However, due to the complexity of this modification and the only minor performance benefits, we do not think that these optional airflow pathway modifications are necessary.
2. In the case of supply shortage of the respiratory filters, there is a second adapter piece that we have designed to replace the viral filter with two P-100 filters made by 3M (Figure 4a). This adapter interfaces with the snorkel-to-ISO part, and can be added onto the prototype. The input port is a male ISO standard 22mm. The output port is designed to connect to standard cartridge filters by 3M. Two gaskets are required to mount these filters (Figure 4b), and typically must be ordered separately. The validity of using P100 industrial filters for infection control is first provided by Gardner et al [15], in which they tested the viral penetration through an N95 and P100 respirator, and showed that those respirators filtered out viral particles at claimed filtering efficiencies.

**Figure 3.**
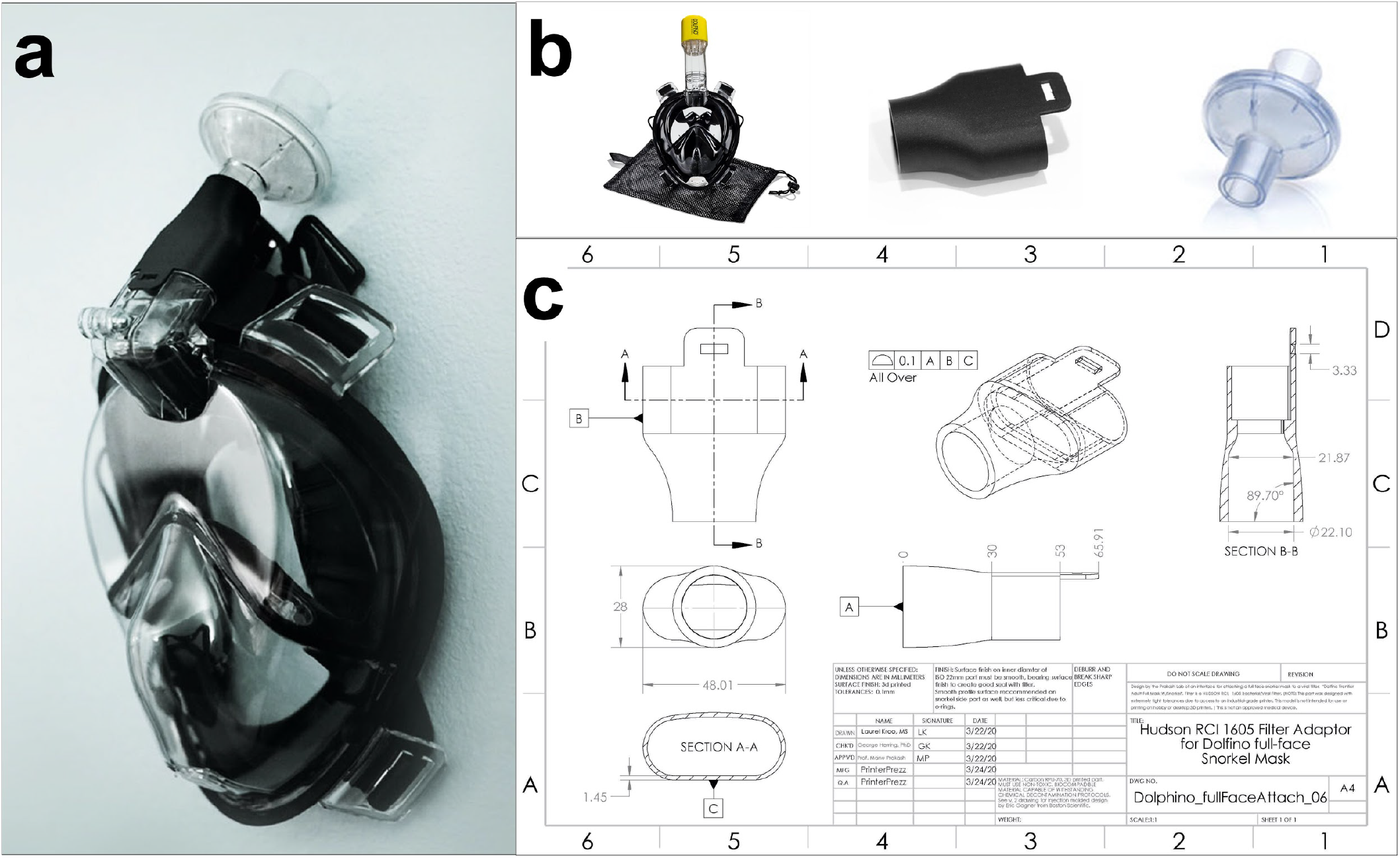
Figure 3: (A) This is the first solution that enables use of a snorkel mask to be used as a full reusable PPE; including (B, from left to right) Dolfino full face snorkel mask, an adapter, and a Hudson RCI 1605 inline filter. (C) The design is based on the connector standard ISO 5356-1:2015 Anaesthetic and respiratory equipment - Conical connectors - part 1: Cones and Sockets). A newer version of this part was adapted for use with injection molding by Boston Scientific.

**Figure 4.**
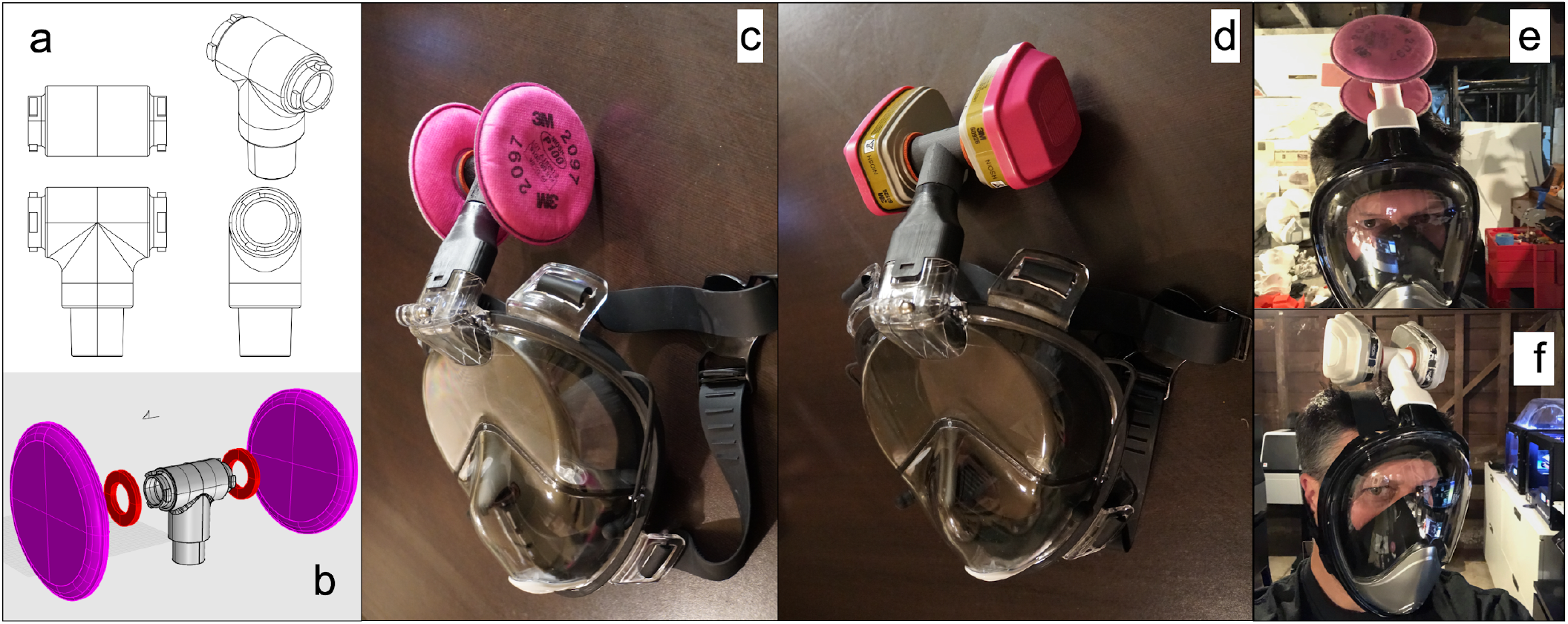
(a) Part Design of a male 22mm ISO to two standard turn socket ports for industrial-grade filters. (b) Assembly with two P100 filters (Part 2097, 3M), (c-d) Mask assembly with multiple types of industrial NIOSH filters. (e-f) Full assembly on Dominic Peralta with different filter types. A list of possible industrial filters that can be potentially used are provided in Appendix A Table 4.

### 2.5 Pneumask-S: Surgeon-Specific Design

The specific needs of hospital personnel that are required to perform ‘sterile-field’ procedures was recognized as an important, separate challenge with regards to design. During surgical procedures, the air exhaled by the surgeon must be directed away from the patient, and preferably be filtered [16]. Pneumask-S can also be used for surgeons who fail N95 fit tests, as typical PPE alternatives, such as positive air pressure respirators (PAPR), are unsuitable while operating over a surgical field. The circumstance of a contagious/infected patient posing risk to hospital personnel in a surgical environment (sterile-field) poses a unique occupational health challenge at the current time of publication.

The chin valve can be modified to be permanently closed/blocked for this prototype (Figure 2D), such that all exhaled air is ported up through the two outside channels to the top of the snorkel. A single-part design has been developed for connecting the center channel of the snorkel (for inhale) to a male filter port, and the 2 outside channels (for exhale) to a separate male filter port. This design has the benefit of fully separating the input and output streams, while avoiding use of the chin valve. A one-way valve on the input prevents any exhaled air from going back to the filter, and lowers fogging and CO2 buildup (optional). The exhale port needs to be connected to a one-way valve (mandatory).

## 3 Testing and Ongoing Clinical Validation

The primary function of respiratory PPE is to protect the wearer from exposure to pollutants present in air, specifically in this case from particles exhaled/sneezed/coughed by an infected individual. The residual exposure of the wearer depends on three independent additive components: the leak at the face, the penetration through the filter and the internal contamination.

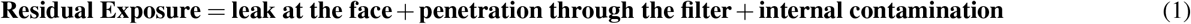

The leak at the face depends on how well the mask forms a seal with the wearer face, or said differently, how well the mask fits the wearer face. The fit is clearly dependent on the mask shape and the morphology of the individual face, and should be determined systematically for each wearer with each of the mask models used. As the origin of the leaks is a breakthrough in the sealing, the fit is considered independent from the nature of the pollutant.

The penetration through the filter depends on the efficiency of the filtering material to remove particles from the air. The filtration efficiency is clearly a characteristic of the filter material and the nature of the particle, and is independent from the individual and from the mask shape. The filtration efficiency has to be measured under normalized conditions, or at least with well characterized particles corresponding to the pollutant the wearer has to be protected from. Filters with less than 99% filtration efficiency at inhalation flow rates will interfere with fit tests and can result in a false fit test failure. This problem can be addressed, in quantitative fit testing, by testing masks with lower filtration efficiencies in the N95 mode available in some TSI Portacount machines.

The internal contamination is mainly due to an inadequate maintenance of the mask, but could be significantly reduced by wearer training, adapted maintenance, and storage protocols (such as sterilization). Currently, the particles exhaled by the mask wearer have the same size range as those generated by the patients, and which the mask should protect against. The global protection factor of the mask cannot be determined while the mask is being worn, as the wearer-exhaled particles could be misinterpreted as a “leak”.

**Thus, for multiple scientific reasons, the fit and the filtration efficiency must be determined separately**. The exposure of the wearer will be considered adequate when both fit and filtration efficiency criteria are respected.

### 3.1 Fit Testing and Seal

For their intended purpose, the seal tests of snorkel masks are done by the manufacturing companies underwater. However, the sealing ability of the snorkel masks on dry skin is unknown. Per CDC and NIOSH regulations on the use of elastomeric respirators, a fit test can be performed in the same manner as N95 respirators to ensure seal and safety to use for an individual [19]. At this time, we recommend that all practitioners seeking to utilize these masks perform a fit test under standard N95 fit test conditions.

In addition to this recommendation, additional fit test experiments have been performed in our laboratories. Practically, two types of fit test can be conducted:

- Qualitative fit test: a liquid aerosol with a sweaty or bitter taste are generated within a confinement around the head of the mask wearer. The result of the fit test is based on the detection of the taste under the mask.
- Quantitative fit test: the method is based on a particle counting outside and inside the mask in parallel using the TSI Portacount device. The ratio out/in gives the fit factor.

#### 3.1.1 Qualitative Fit Test

At the University of Utah, we performed a qualitative fit test on 3 separate volunteers, utilizing our 3D printed adapter and both an HME anesthesia circuit filter and a HEPA anesthesia circuit filter on a Dolfino Frontier mask (Figure 5). Out of 3 volunteers, 2 were male and 1 was female, and both males had failed their fit test in the past using regular N95 respirators. The fit test was performed by the standard University of Utah Operating Room team as for N95 tests, as part of the emergency COVID-19 response in order to evaluate emergency countermeasure personal protective equipment. Importantly, all 3 individuals passed their fit tests. This preliminary result seems to indicate that the fit seal satisfies the minimum requirements for an N95 respirator or elastomeric respirator.

**Figure 5.**
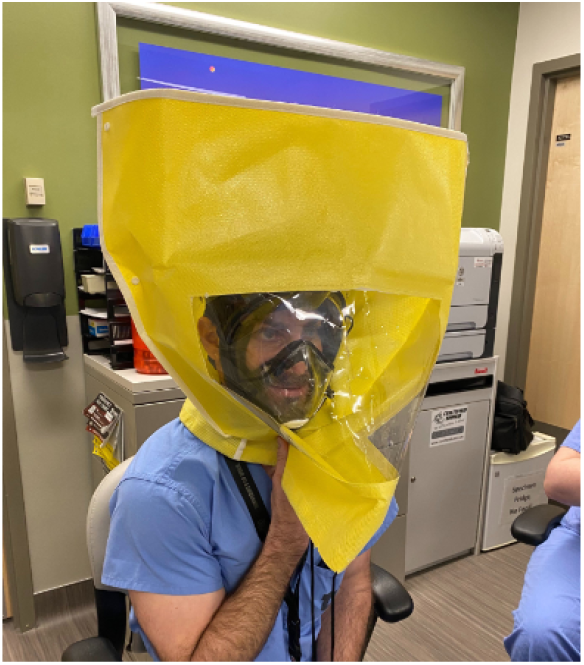
A qualitative fit test was performed by the standard University of Utah Operating Room team using the same protocol suggested by CDC and NIOSH on elastomeric respirators and N95 masks.

**Figure 6.**
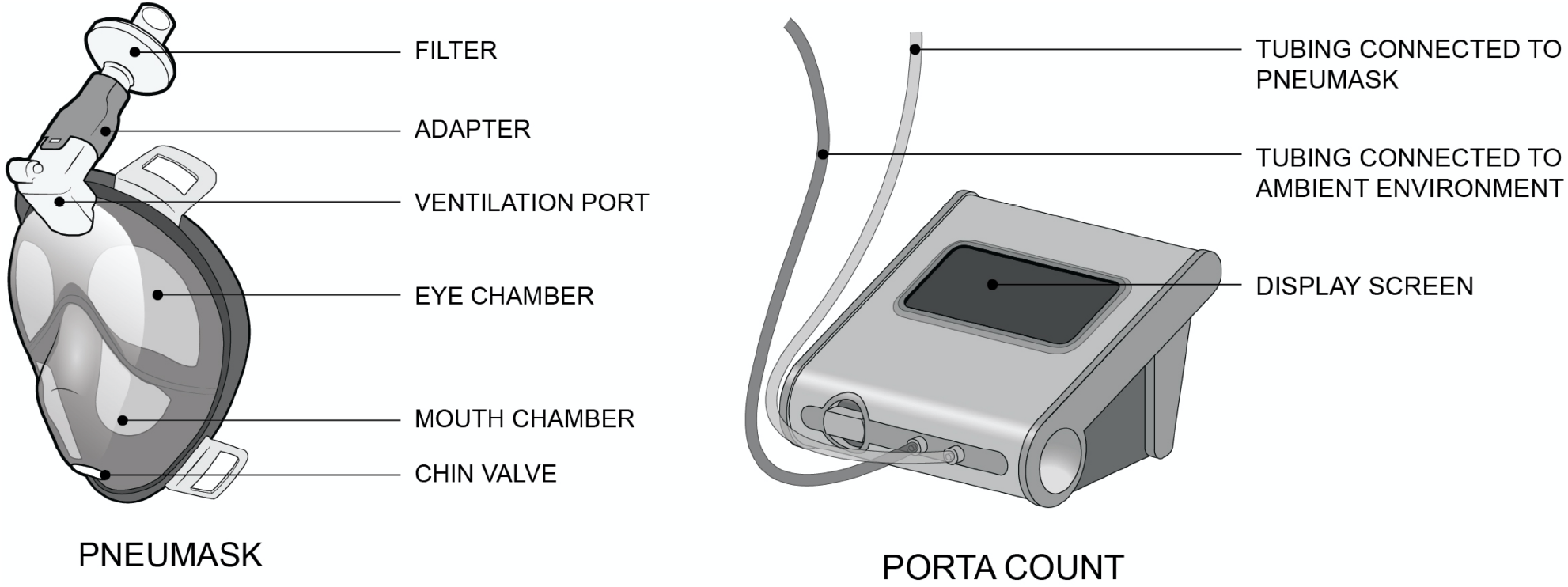
Pneumask and PortaCount essential components.

**Figure 7.**
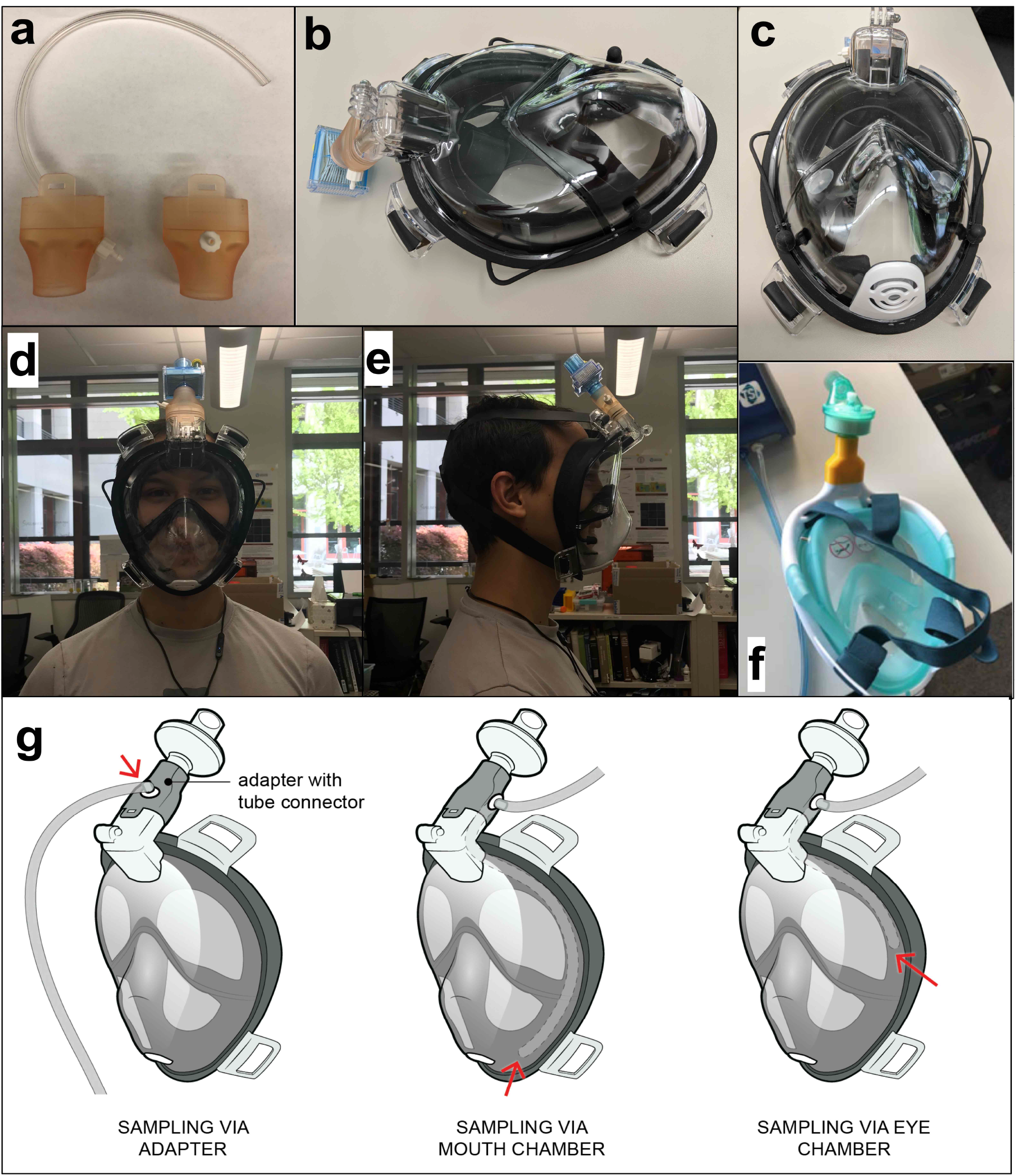
(A-B) Modification of adapter for quantitative fit test. The sampling of the air inside the mouth chamber is achieved by a flexible tubing running through the side channel and connected to the sampling port on the adapter. This method avoids possible deconstructions on the mask surface. (C) The yellow arrow points out the tip ending of the sampling tube. (D) The front view and (E) side view of our testing subject wearing the mask. (F) Subea Decathlon mask used for quantitative fit testing at EPFL. (g) Different testing methods, depicting measuring the exact position for Portacount sampling tube. Red arrows point to the tube inlet position inside the mask.

#### 3.1.2 Quantitative Fit Test

The fit test is only meant to measure the ability of the mask to form a seal with the wearer’s face, and not the efficiency of the respiratory protection. The principle of this test consists of successive measurements of the particulates concentrations inside and outside of the mask during normalized exercises. The ratio between the external and the internal concentrations is called fit factor (FF). The relevancy of the results is dependent on a few assumption:

- The efficiency of the filter is high enough to assure that the particle penetration rate is insignificant compared to the expected leak rate, within the range of the measured particle size (0.015−1 *µ*m). P3, N100 or HEPA filters are generally required to reach this specification (filtration rate *>* 99.95 % at 0.3 *μ*m), with a theoretical FF *>* 2000 in case of perfect fit. For filters with significant penetration rate (P2 or N95), the measure is based on a smaller subset of particle sizes (around 0.04 *μ*m) using the N95 protocol to avoid the counting of the filter-penetrating particles.
- The range of the particle size measures by the Portacount (0.015−1 *μ*m) has been selected to stay mostly outside of the range of particulates generated by human exhalation, apart from smokers (it is inadvisable for smokers to perform these tests, but at a minimum, they should not have smoked within 30 minutes of testing). This is necessary because the particulates generated by mask wearer would otherwise be misinterpreted as a leak into the mask.
- The ambient particle count exterior to the mask, in the particle size range measured by the machine, must be significantly higher than the quantity of particles that could be generated by the wearer by any method. Quantitative fit testing units such as the TSI Portacount are programmed to abort testing if the ambient particle count decreases under a minimum level.

Practically, the quantitative fit test will not measure only the leaks at the wearer’s face, but also any leaks in relation with the connection after the filter or with the exhaust valve. In this way, high fit factors reflect both that the leakage at the wearer’s face is acceptably low (wearer dependent), but also that the residual leaks at the level of the adapter and the chin valve are acceptable as well (wearer independent).

Here we present quantitative fit test results. We will first discuss the multiple tests run at Stanford University (through multiple groups) on the Dolfino Frontier Mask. We will then review the results both from Stanford and from EPFL on the Subea Decathlon mask. Both mask models passed quantitative fit testing with fit factors that meet the industry-standard threshold for typical half-face elastomeric respirators.

##### Stanford Prakash Lab Testing: Experiment Parameters

- Fit measured using a Portacount Pro+
- OSHA Standard
- Half Mask protocol (non-N95)
- 4 candles to provide a constant ambient particle source
- Test Subject: 28 year old male, no facial hair, no history of smoking
- HME HEPA filter (Pall Ultipor 25)
- Snorkel Mask (Dolphino or Decathlon)
- Modified adapter from Formlabs high temperature resin

In order to collect a thorough measurement, each test was repeated in three locations, sampling from the mouth chamber, the eye chamber, and directly from the adapter, right after the filter. This required building two modified adapters. The adapter which sampled directly after the filter was modified by drilling a hole in the adapter between the filter connection port and the mask port. The drilled hole was cleaned and smoothed before a luer lock connector was press fit into the hole and sealed using epoxy. The epoxy was allowed to cure for 24 hours and the assembly was subsequently washed with isopropyl alcohol for 2 minutes. The adapter which sampled from the eye chamber and mouth chamber used a flexible tube to sample air from a desired location. This adapter was modified by drilling a hole large enough for the flexible tube to pass through. The hole was once again cleaned and a luer lock connector was pressed into the tube. The luer lock connector and tube were press fit into the drilled hole and sealed using epoxy. The epoxy was allowed to set for 24 hours before the entire assembly was washed for 2 minutes in isopropyl alcohol. Both adapters were allowed to dry completely before any tests were conducted.

##### Dolfino Frontier Mask Experimental Results

Results from the Stanford Prakah Lab are posted here for the fit results on the Dolfino Frontier mask. Further tests were completed independently at Stanford Occupational Health and Safety which confirmed that the Dolphino mask passes the quantitative fit test.

Two additional fit tests were conducted at Stanford Environmental, Health and Safety – one completed using the requirement of a half-face elastomeric respirator and another using the fit factor for a full-face tight fitting Air Purifying Respirator, and the results are shown in Figure 10. The participant (female) wearing Dolfino Frontier with a custom adapter, and a Pall Ultipor 25 breathing filter, who is typically a size M, was still able to pass the quantitative fit test well beyond the requirements on both tests. The minimum passing fit factor was 100 for half-face respirator and 500 for a full-face respirator. The activities that were tested while wearing the mask included bending over (50 seconds), jogging in place (30 seconds), moving head side to side (30 seconds), and moving head up and down (30 seconds), and the fit factor outcomes were all above 750. Please note that, although the test yielded positive results, this was conducted with a limited testing sample and does not yet indicate any certification/endorsement from EH&S of the product. Each entity should also conduct its own evaluation and testing before use.

**Figure 8.**
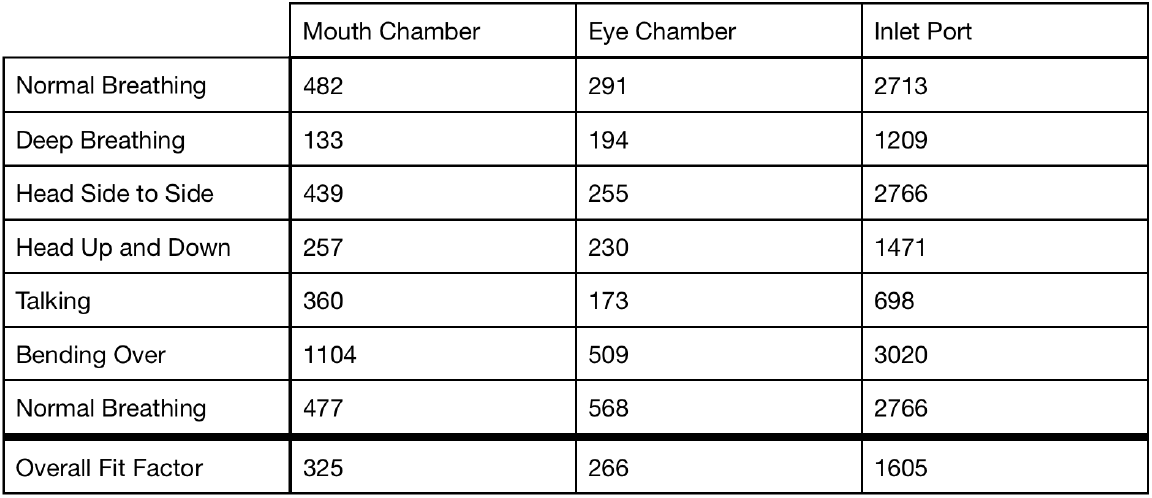
Fit factor results when the Portacount sample tube was connected to the mouth chamber, eye chamber, and inlet port of a Pneumask. This mask consisted of a Dophino mask connected, via an adapter, to a HEPA rated mechanical HME filter (Pall Ultipor 25). These tests were completed in half-face respirator mode and the mask passed in all three cases.

**Figure 9.**
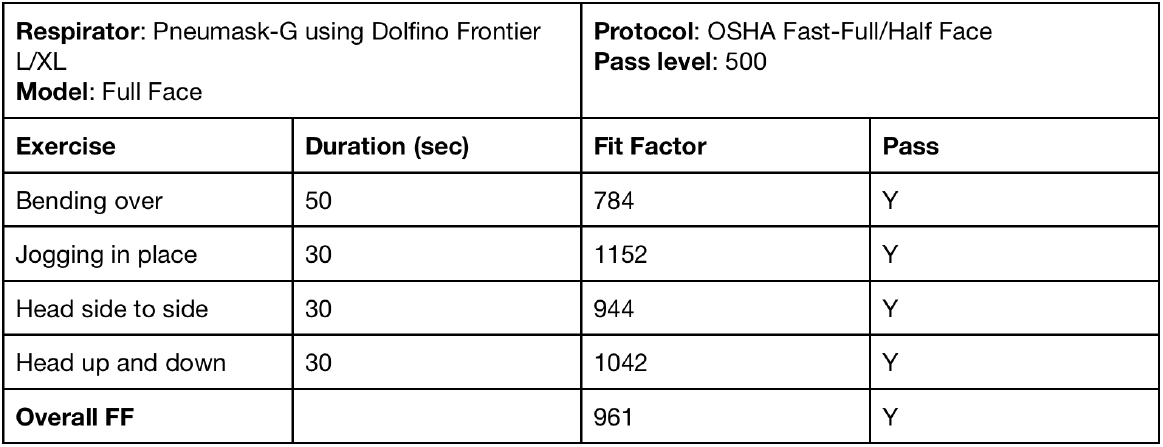
Results of quantitative fit tests using PortaCount 8048, conducted by the EHS team at Stanford Health Care. The sample is obtained from the adapter site, using a modified adapter as shown in Figure 7f.

**Figure 10.**
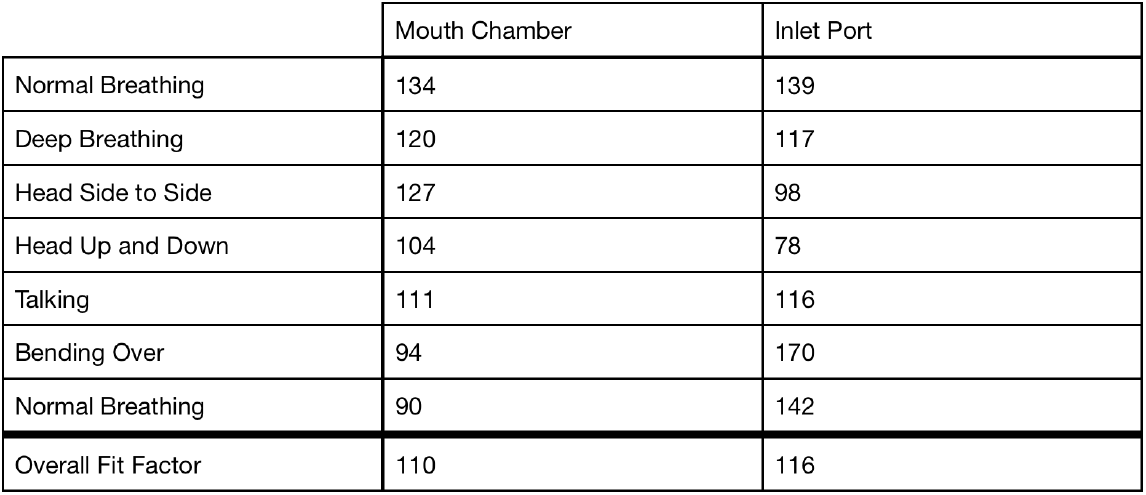
Fit factor results when the Portacount sample tube was connected to the mouth chamber and inlet port of a Pneumask. This mask consisted of a Decathlon mask connected, via an adapter, to a HEPA rated mechanical HME filter (Pall Ultipor 25). These tests were completed in half-face respirator mode and the mask passed in all cases.

##### Subea Decathlon Experimental Results

Testing was undertaken both at Stanford University and at EPFL with quantitative fit testing for the Subea Decathlon Mask. Below are the results from our tests at Stanford. Important Note: The adapters used in the above test did not fit the Subea mask as well as the Dolphino mask. Tape had to be used to form a better seal. Thus these results should be interpreted as a lower bound on the sealing capabilities of the mask.

Validation of the fit of the Decathlon mask was completed in a separate series of experiments at EPFL, also using a Portacount Pro+ in N95 mode and following the OSHA 29CFR1910.134 protocol. The Decathlon Easybreath mask was connected to a medical grade HME filter (DAR adult-pediatric electrostatic filter HME, small) with a 3D-printed PLA connector. The mask was in Pneumask-G configuration (3 snorkel ports connected to the filter, chin valve non modified). The mask was connected through the silicon skirt of the eyes chamber, as indicated in Figure 7f, using the standard connector sold by the manufacturer of the particle counter. In N95 mode, the test was positive for the two individuals (men, freshly shaved) tested, with a fit factor of 200+, which is higher than the requirement for half-masks (100). Removing the chamber valves to connect permanently the eye chamber and the mouth chamber led to the same results. This test was run in N95 mode because the test was completed using an HME filter which was not HEPA rated (fit factors around 4 would have been obtained with the N100 normal protocol under the same testing conditions). The full results from EPFL testing are shown in Figure 11.

**Figure 11.**
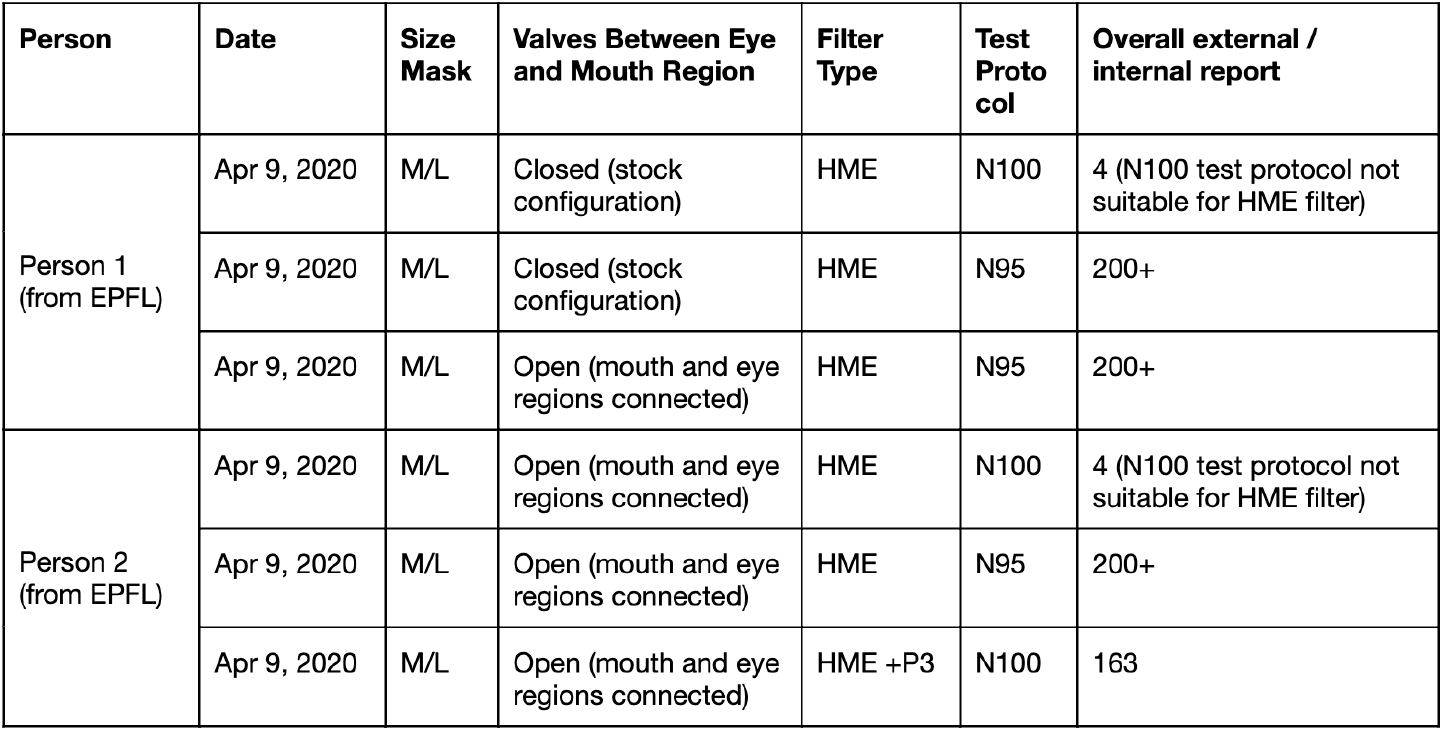
Fit factor test results from EPFL (translated from French). Note that we have identified a common testing issue mistake – you cannot use the default N100 mode unless the respirator or HME filter is rated above 99% at respiratory flow rates.

**Figure 12.**
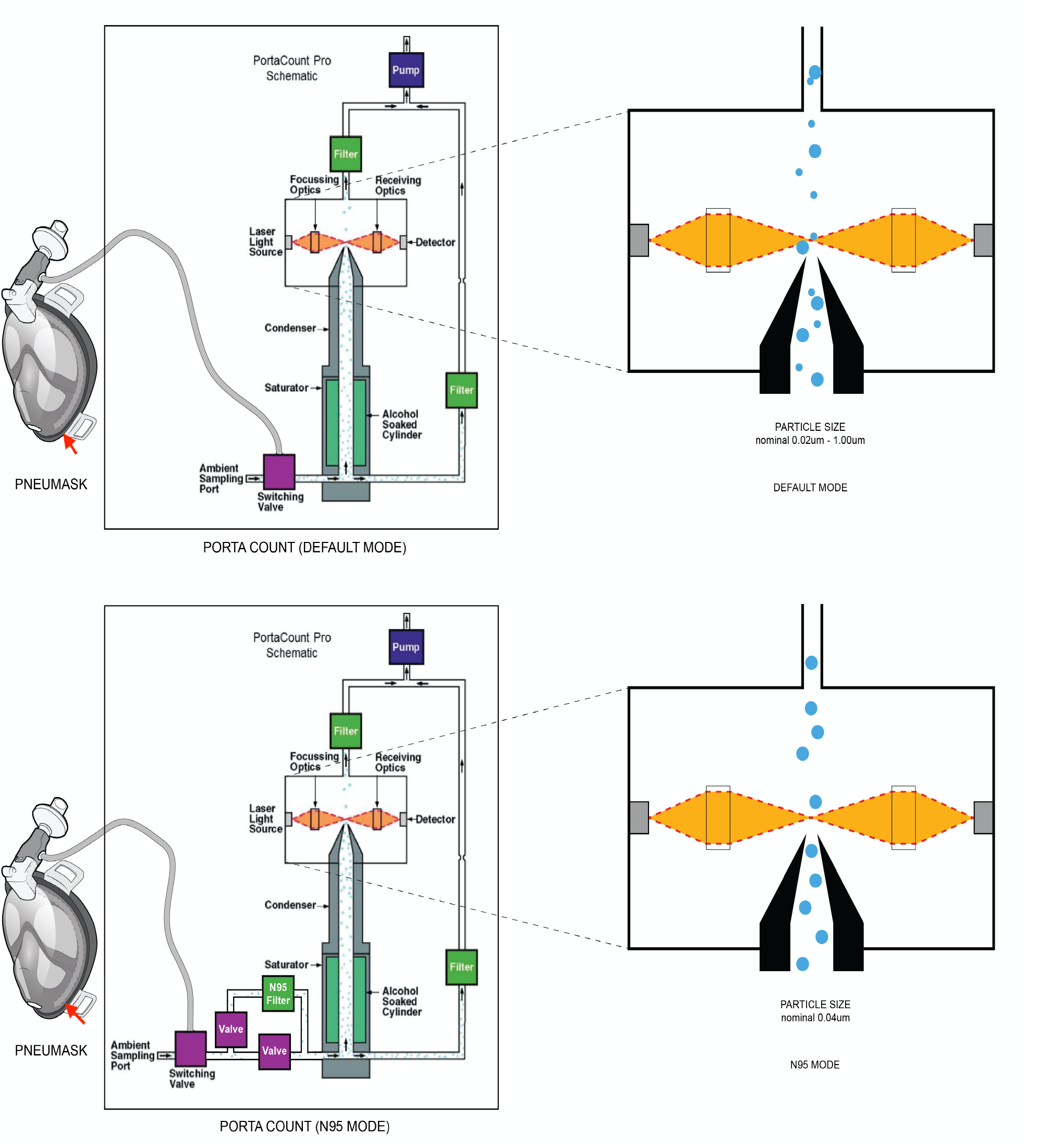
Difference in PortaCount’s functionality across the default and N95 modes.

**Figure 13.**
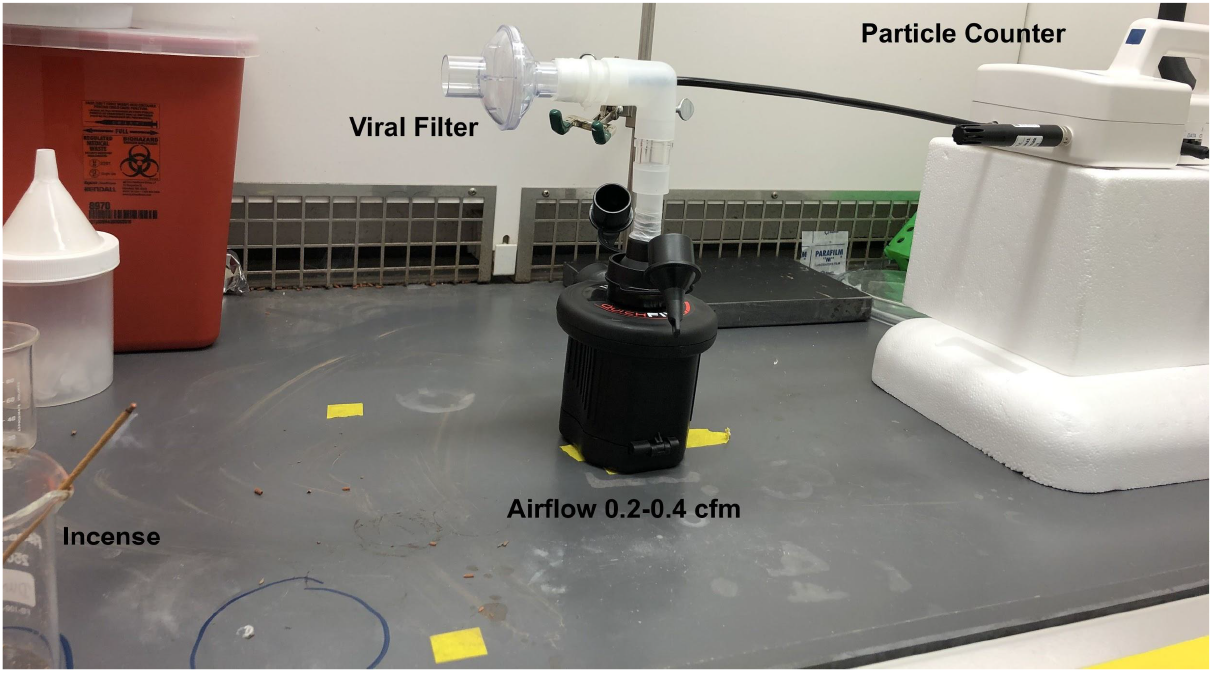
Setup for measuring filter efficiency.

##### Summary of Fit-Test Findings

The successful results for the fit test with the different individuals used in this study indicate that the Dolphino and Decathlon masks both form acceptable seals, showing also that the custom adapter and the chin valve do not generate significant leaks. The measured fit factors correspond to the requirement for elastomeric half-mask. The remaining performance of the mask depends on the efficiency of the filter, which is attached to the mask.

The position of the sampling point gives similar results between the mouth and the eyes chambers. However, a sampling point directly connected on the custom adapter shows significant higher fit factors, which seem over evaluated, probably due to the proximity of the filter. In this case, the measured particle concentration should not be relevant of the real concentration in the breathed air.

The use of the N95 protocol of the Portacount is important for the fit evaluation when HEPA filters are not available, especially with HME virus filters. The efficiency of the filters should be measured independently of the Portacount system to assure safe working conditions.

### 3.2 Filtration Efficiency Testing

We developed an simple experimental test rig and method for testing the particle filtration efficiency of various materials. Please note this setup is not the standard testing method which typically uses the TSI Automated filter tester 8130A. The setup pictured in figure 14 includes a LightHouse handheld particle counter (Model 3016 IAQ), Intex QuickFil 6C Battery Pump, a rubber stopper with 2 holes covered by 2 kim wipes to mitigate the airflow, Incense: Satya Sai Baba Nag Champa 100 Gram, connectors (universal cuff adaptor, teleflex multi-adaptor), and filters to test (Hudson RCI Main Flow Bacterial/Viral Filter, Romsons HME Disposable Bacterial Viral Filter, Pall Ultipor 25 filter). The pump with the rubber stopper, covered by 2 kim wipes, in it, provides an airflow within a range of 5.6 - 11.32 l/min to mimic that of breathing. The incense produces particles of various sizes, including those in the range picked up by the detector (0.3 *ε*m - 10 *ε*m). With the pump on, we measure the number of particles produced by the incense. Then we place the filter on the setup and run the particle counter to measure the number of unfiltered particles. To calculate the filtration efficiency, we calculate the ratio of unfiltered particles to the number of particles produced by the incense, and then subtract from one. The filter efficiencies for the 3 filters tested are reported in Figure 15.

**Figure 14.**
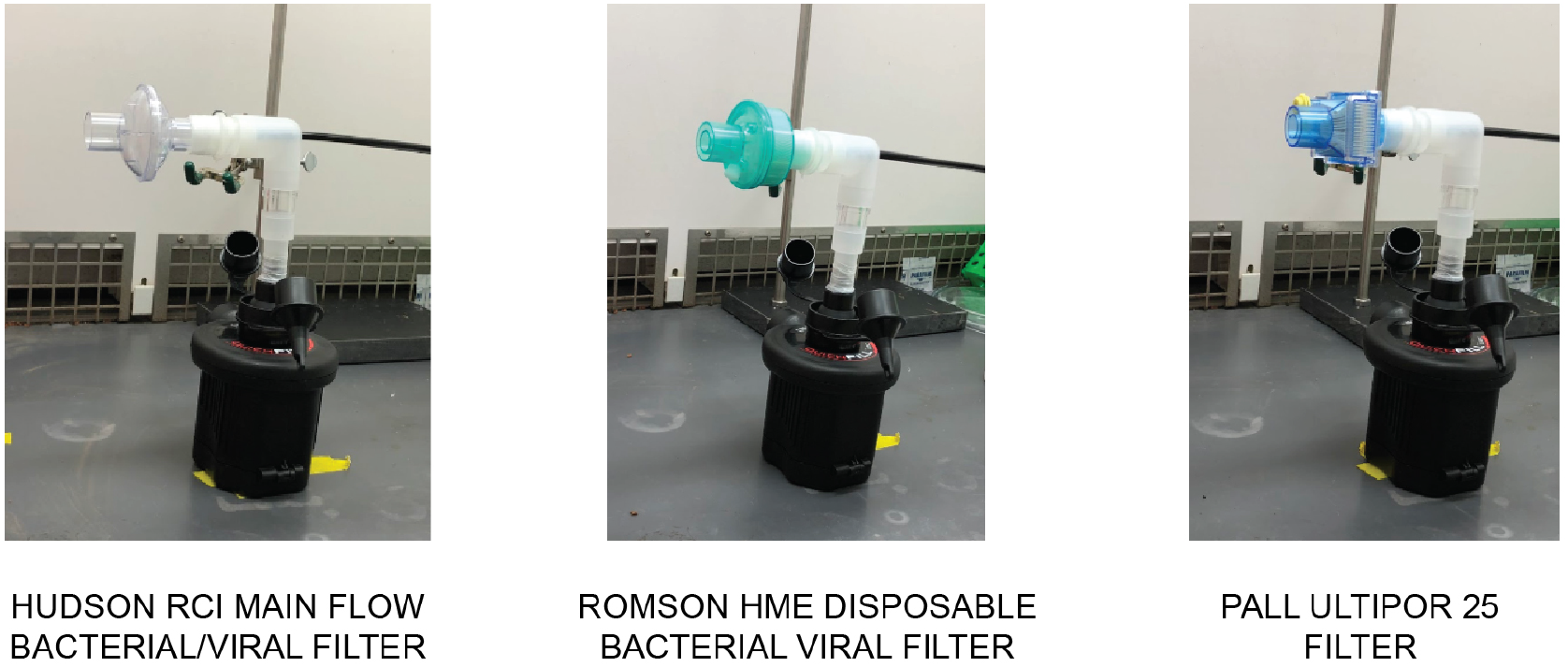
Variety of filters tested.

**Figure 15.**
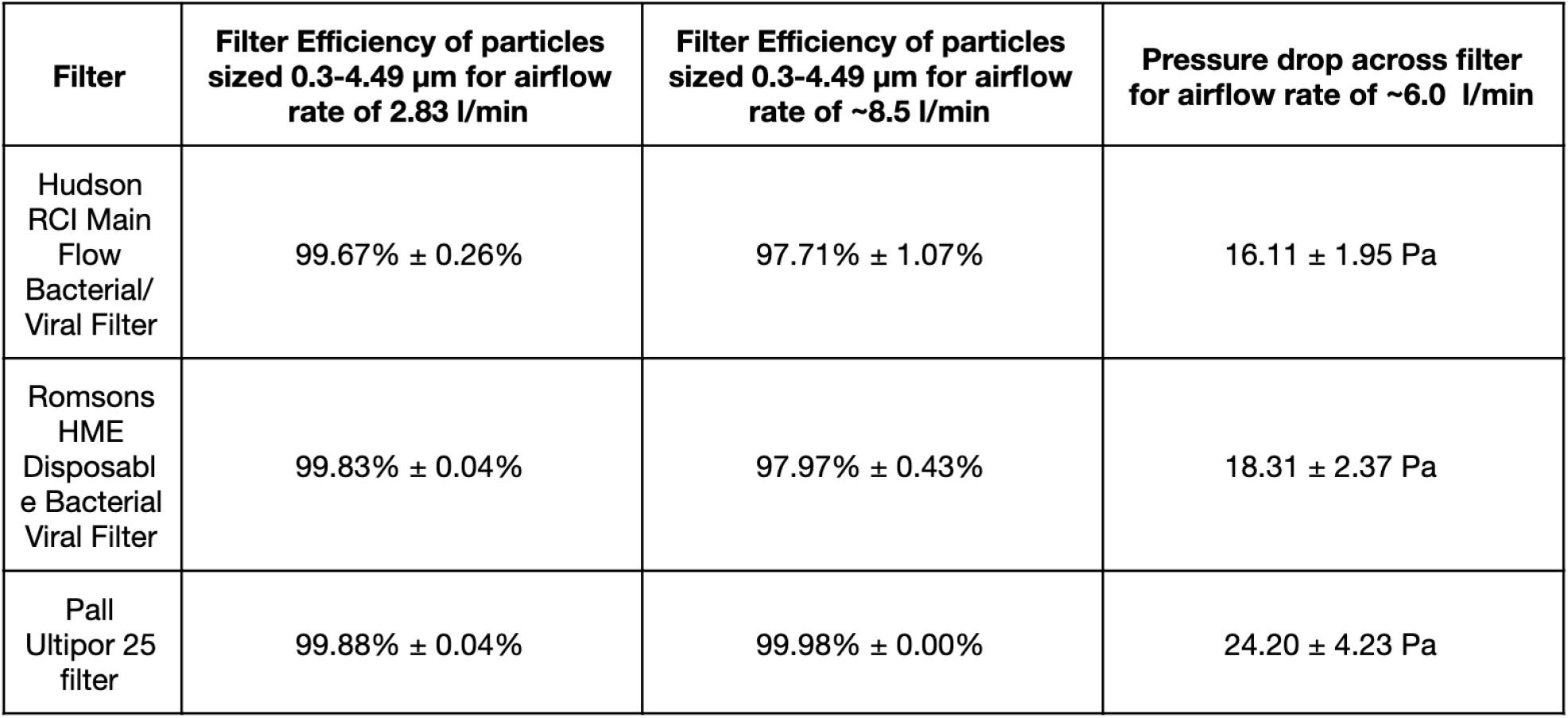
Filter efficiency and pressure drop across filters.

We constructed an experimental system for measuring the pressure drop across various materials, including N95 masks, during inhalation and exhalation. The setup in picture Figure 16 includes an Intex QuickFil 6C Battery Pump, a Honeywell AWM700 Airflow sensor, a Honeywell ABPDLNN100MG2A3 Pressure Sensor,a rubber stopper with 2 holes covered by 2 kim wipes to mitigate the airflow, connectors (universal cuff adaptor, teleflex multi-adaptor), and filters to test (Hudson RCI Main Flow Bacterial/Viral Filter, Romsons HME Disposable Bacterial Viral Filter, Pall Ultipor 25 filter). The pump with the rubber stopper, covered by 2 kim wipes, provides an inhalation or exhalation airflow within a range of 0.2-0.4 cfm to mimic that of breathing. With the pump on, we measure the airflow applied to the mask, and the differential pressure drop across the mask. The pressure drops for the 3 filters tested are reported in Figure 15.

**Figure 16.**
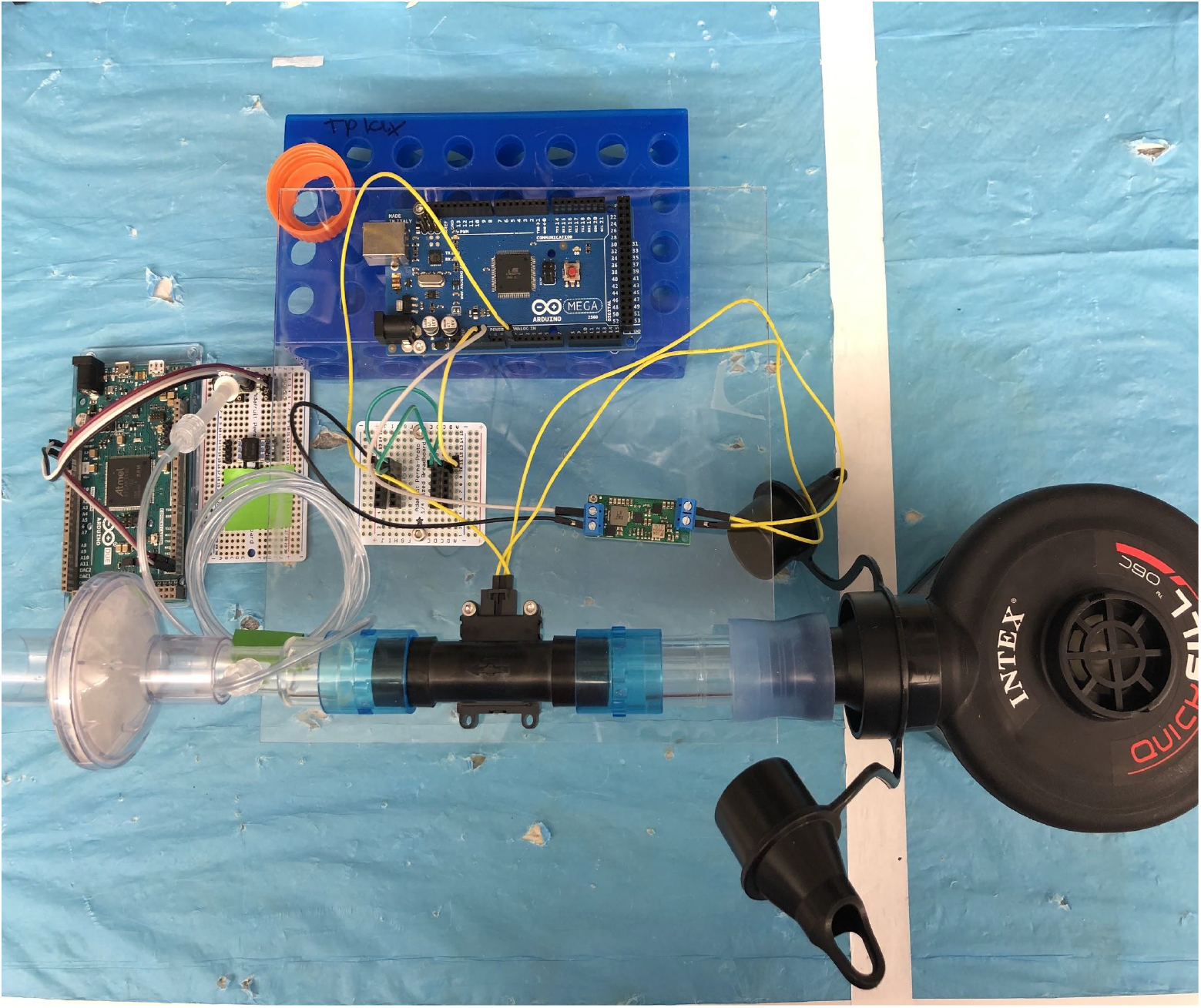
Setup for measuring pressure drop across filter.

#### 3.2.1 Summary of Filtration Testing Findings

We have found that the Decathlon Subea masks and the Dolphino masks are both capable of forming a seal that exceeds the standards required for half-face respirators and N95 masks (fit factor >100). The masks must still be properly secured and sized appropriately for the wearer with the fit verified according to the standards of the institution where the PPE is being used. The sealing capabilities of these masks were tested using a TSI Portacount Pro+ (in half-face mode using the OSHA standard) on a system that consisted of the mask, a custom adapter to connect the mask and filter, and a HEPA rated HME filter. The custom adapter was modified to include a sampling port to which the TSI Portacount Pro+ could be connected, allowing measurements at three different places inside of the mask (next to the filter, in the eye space, and in the mouth space). The particle concentrations in all parts of both masks were found to be less than 1 part in 100 relative to the ambient particle concentration (Fit factor of >100). The Decathlon Subea mask was also tested at EPFL, again with a TSI Portacount Pro+, on a system consisting of a Decathlon mask, a custom adapter, and an electrostatic HME filter. The test was run by connecting the Portacount Pro+ to a port which was installed in the rubber siding of the mask next to the eye chamber. The test was run using an OSHA standard in N95 mode (as required by the Portacount Pro+ for filters with <99% efficiency). The Portacount Pro+ reported a fit factor of >200 (a particle count of less than 1 part in 200 relative to ambient conditions) for two different wearers. Repeating the test with the eye chamber and mouth chamber directly connected to allow bidirectional airflow between the two chamber resulted in the same fit factor. The sealing capability of both the Dolphino and Decathlon masks has been shown to exceed the standards for half-face respirators and N95 respirators. These tests were verified at multiple locations within Stanford and at EPFL using different masks, wearers, adapters, filters, Portacount machines, and machine operators.

### 3.3 Evaluation of the Chin Valve

#### 3.3.1 Exhalation Valve Leakage Test

One concern our team had was the ability for particulates to enter through the chin exhalation valve. As these snorkel valves are not medically approved, it is necessary to understand the performance of the valves to assess safety. The NIOSH standard “Exhalation Valve Leakage Test” (Section 84.182) stipulates the pressures and flow rates necessary for equivalent performance standard to typical N95 respirators. Particularly, it states that: “(a) Dry exhalation valves and valve seats will be subjected to a suction of 25mm water-column height while in normal operating position. (b) Leakage between the valve and valve seat shall not exceed 30 milliliters per minute.” [28] With this specification in mind, we conducted the following test to determine technical equivalency to NIOSH standards, investigating if this sports equipment is adequate to meet industrial device standards. A chamber was glued around the chin valve using epoxy, and an 8mm tube was connected to this chamber (Figure 17A). The 8mm tube is connected to an opened water tank (Figure 17B). First, the water was allowed to equilibrate by manually opening the chin valve briefly (Figure 17B). Then, the mask was lifted up to create a water column of 30mm (Figure 817C), which corresponds to a negative pressure of 30mmH2O applied to the chin valve. As time passes, the small leakage of the chin valve reduces the negative pressure and therefore the height of the water column. The leak of the chin valve can be approximated as the ratio between the volume of air in 10mm of tube length, and the time it takes for the water column to go from 30mmH2O to 20mmH2O. On average, this time was 21 seconds. As a consequence, in steady state and for a negative pressure of 25mmH2O on average, the leak of the chin valve is on average 1.5mL/min (12 measurements, standard deviation 0.3mL/min). We conclude that the leak flow of the exhale valve (chin valve) is lower than the maximum flow allowed to comply with the NIOSH regulation (30mL/min for 25mmH2O of pressure). It is noted that this test was done on a one specific mask model our team had available, the Decathlon FreeBreath. Although our findings here are reassuring, there is a possibility that different mask models and manufacturers have varying quality of valves. We recommend quantifying this parameter prior to usage (especially prior to large-scale usage of any particular mask brand).

**Figure 17.**
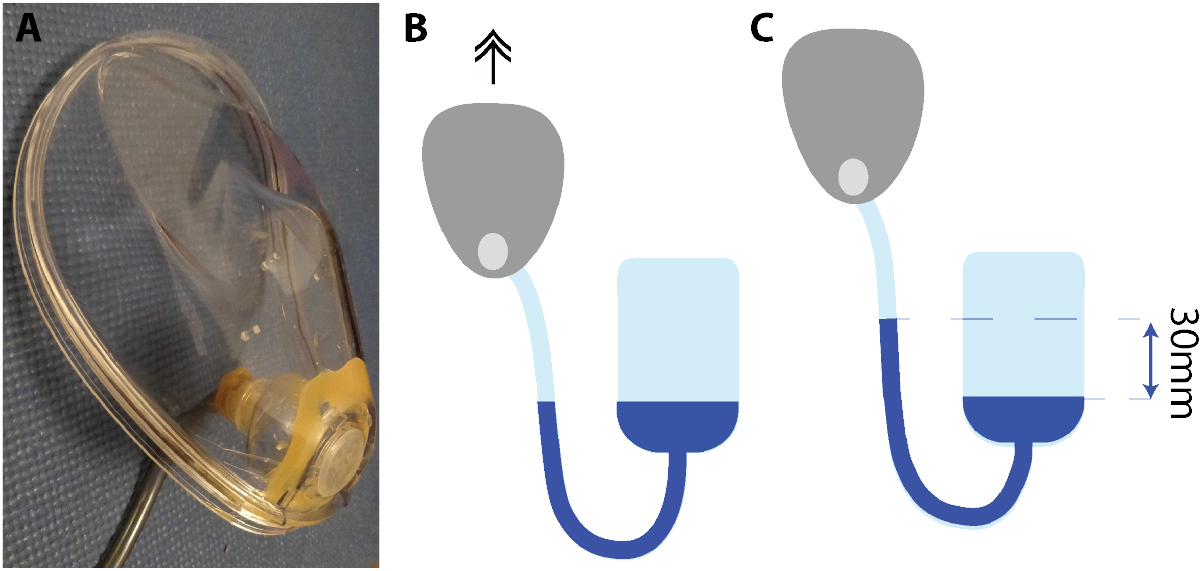
Setup for measuring the leak flow of the chin valve of a Decathlon FreeBreath mask. (A) Connection to an 8mm tube. (B) Connection to an open water tank - system at equilibrium (chin valve opened). (C) The mask is lifted up (chin valve closed) to create a water column of 30mm, which means that a negative pressure of 30mmH2O is applied to the chin valve.

#### 3.3.2 Valve Closure Times Estimation

Aside from the above testing, we have also done the following calculation to see how long the chin valve takes to close (Figure 18A, Figure 19), assuming standard exhale - to assess the likelihood of localized backflow. The following figure (Figure 18B) shows the schematic of a circular chin valve, which is pinned at the center. The valve is assumed to open from the bottom side, moving from vertical position to an angled location after exhalation of air.

**Figure 18.**
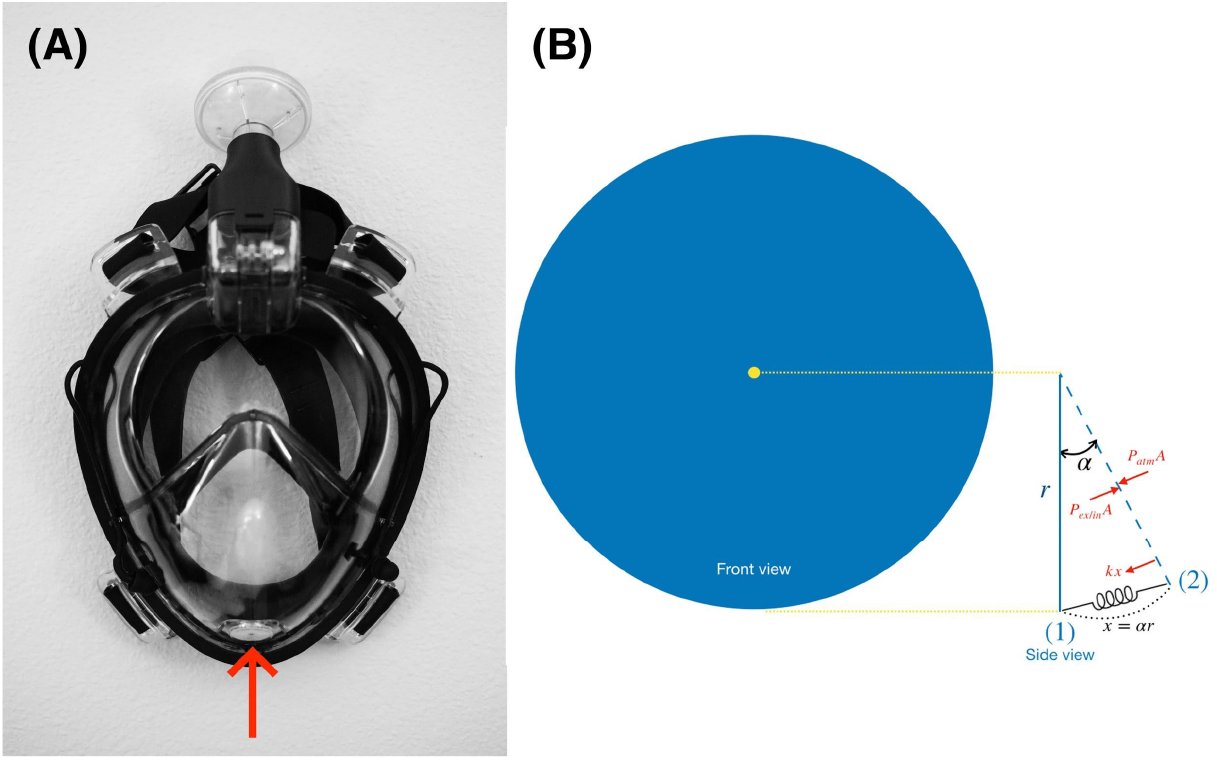
(A) Chin valve on mask, (B) Schematic of a circular chin valve.

**Figure 19.**
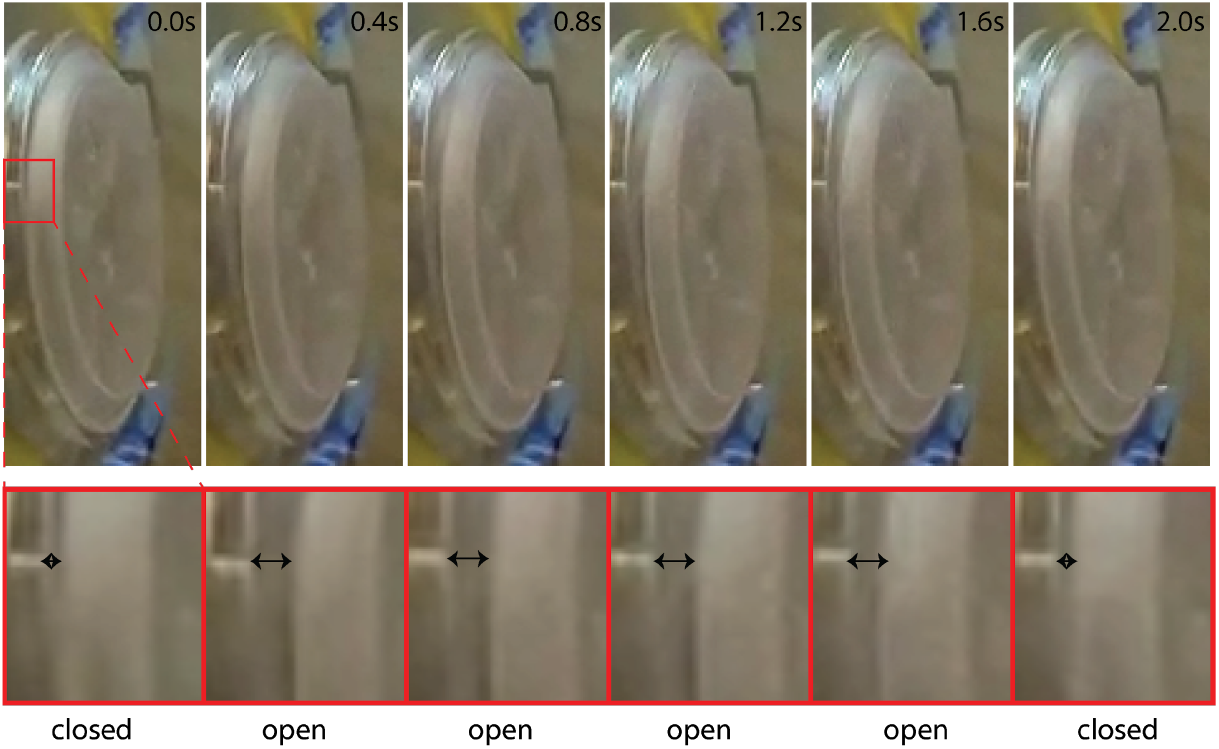
Time-lapse images of the chin valve opening and closing.

**Figure 20.**
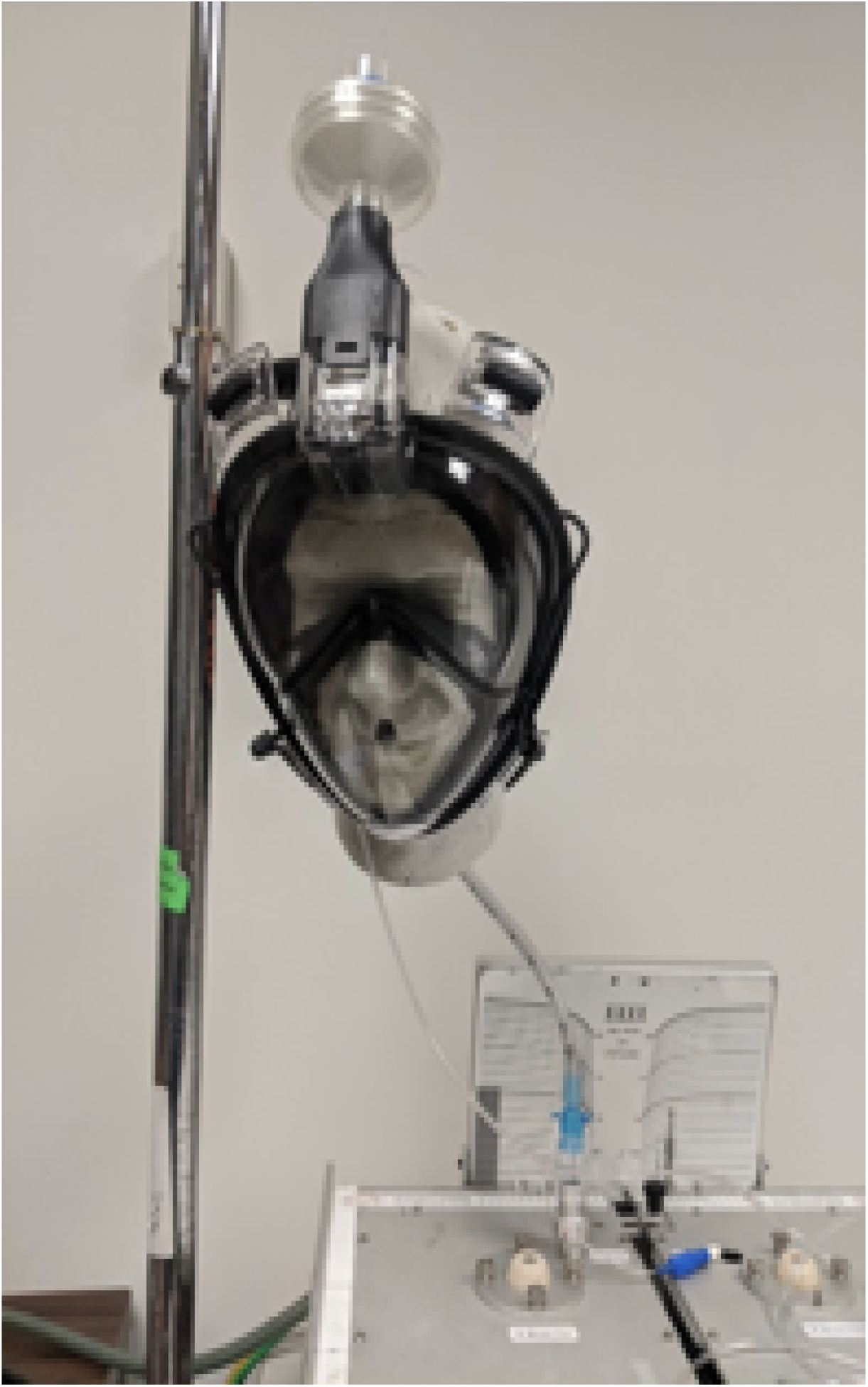
Carbon dioxide washout testing utilizing a headform and simulated lung. Various conditions are being tested to determine the suitability and safety of the proposed snorkel PPE solution in the context of carbon dioxide build up. The snorkel mask used in this setting is Dolfino Frontier.

**Figure 21.**
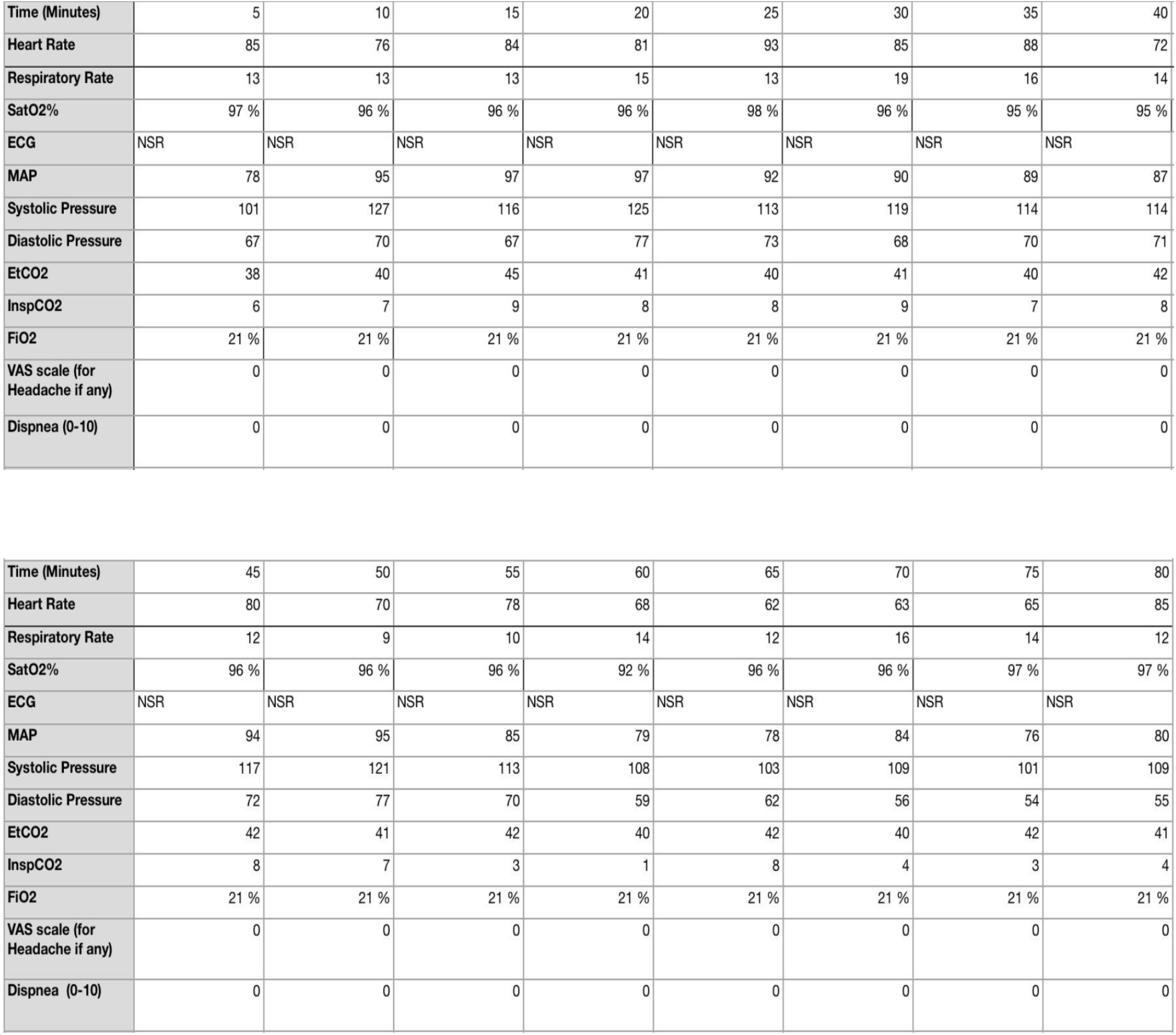
Data from CO2 testing on Dolfino Frontier.

**Figure 22.**
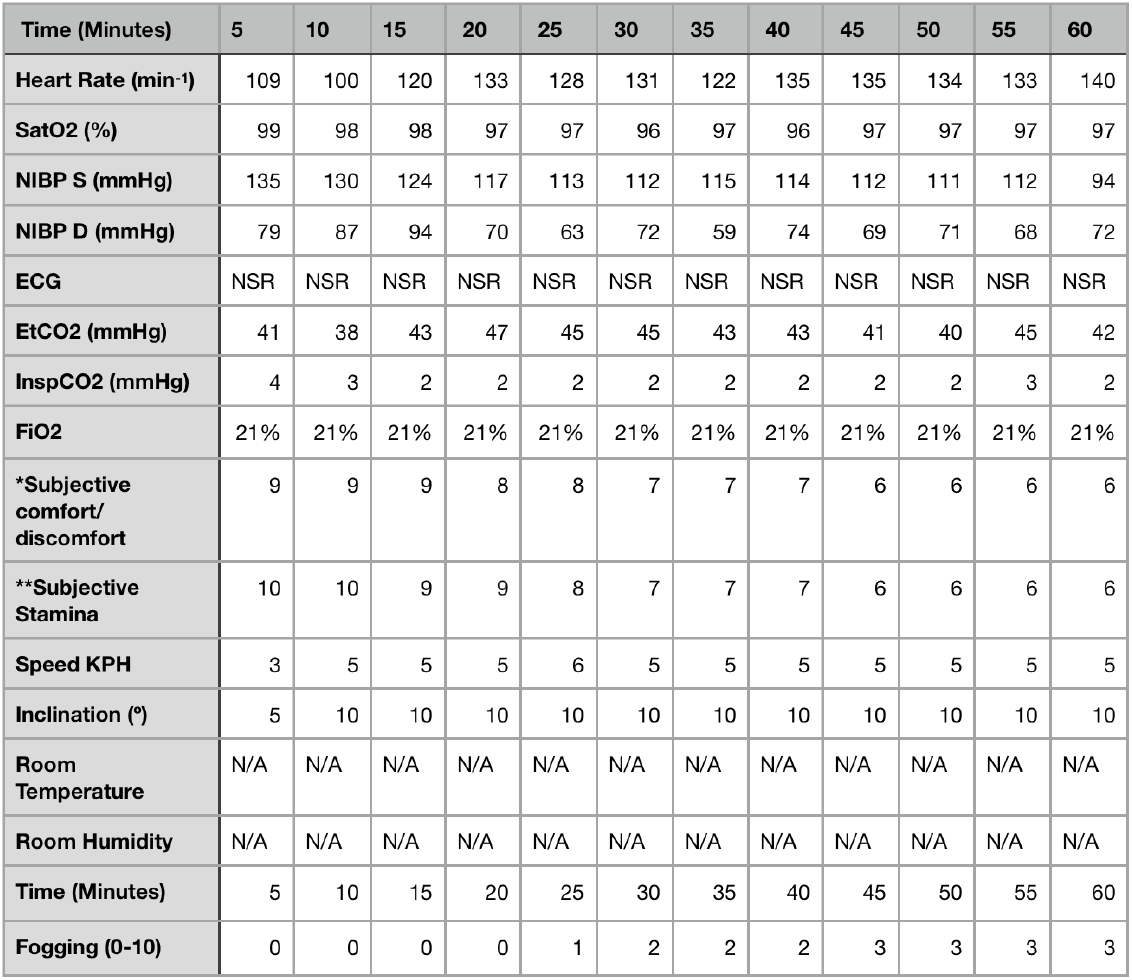
Results of the 1 Hour treadmill test.

Forces acting on the valve include the force due to the gauge exhale pressure, (*P*_*ex,g*_ = *Pex−P*_*atm*_), and elastic forces. The elastic force occurs to return the valve to its original shape, this force is simply modeled by a linear spring formula. These forces should balance for a static valve at (2):

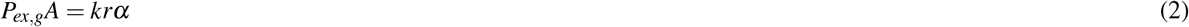

where A and k denote the cross-section area and the elasticity constant of silicon, respectively. Now, let’s assume inhale starts at time t=0 and the valve is in the angled position (2) shown in Figure 18B,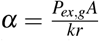

At each time instant, in addition to the elastic force, the pressure forces due to the inhale, *P*_*in*_, and atmospheric pressure are acting on the valve:

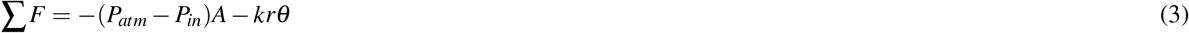

On the other hand:

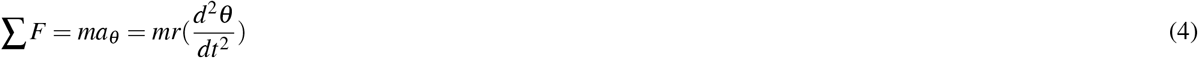

And therefore:

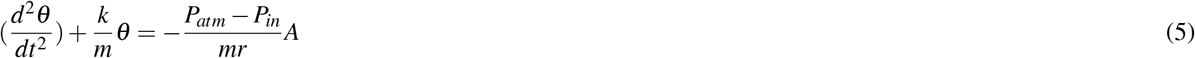

This equation is a second-order ODE with boundary-conditions: *θ* = *α* and 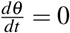 at *t* = 0. This leads to the solution:

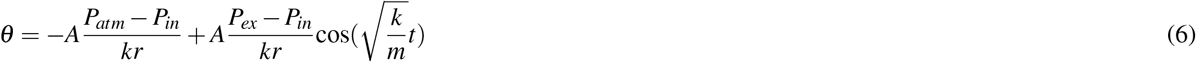

Accordingly, the time it takes for the valve to reach *θ* = 0 equals:

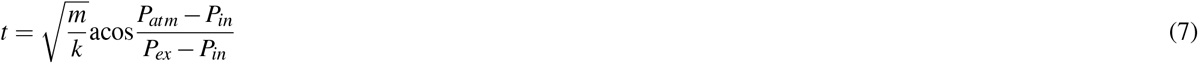

The maximum closure time occurs when we assume there is no force due to pressure during inhalation, *P*_*in*_ = *P*_*atm*_, and equals:

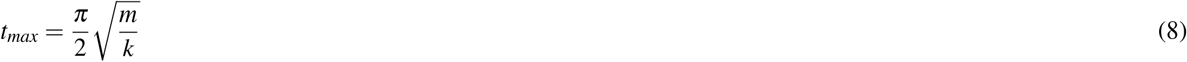

In order to determine a time associated with this dynamics, we need to directly measure some of the properties of materials used in elastomeric valves. For an order of magnitude calculation, we use known numbers; for a silicon valve with density *ρ* = 2.3290g / cm^3^, thickness *l* = 0.35mm, and radius 13.5mm, the mass of the moving section is *m* = 0.233*g*.Assuming elasticity constant *k* = 1N/m, this formula suggests that the valve closes after approximately *t*_*max*_ = 0.024*sec*.

### 3.4 Carbon Dioxide Testing

The accumulation of carbon dioxide in the deadspace is a valid concern that can result in significant risks to healthcare workers utilizing respirators [18]. The accumulation of CO2 can vary greatly in snorkel masks as the connections between the inhalation and exhalation arms, the design of one-way valves, and possibly attached cartridges are very versatile. Also, the issue of CO2 buildup is not unique to full face mask snorkels nor to elastomeric respirators, but is a known factor in the continuous use of disposable N95 respirators as well [19]. For example, Lim et al., 2006 found that up to 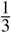 of healthcare workers in the SARS outbreak reported headaches during use of N95 (presumably from hypercapnia) and that 4 hours of continuous use of N95 is associated with headaches [18].

To test this aspect, we utilized the mask-adapter-filter setup attached to a headform and simulated lung (Figure 20) [20, 21]. Carbon dioxide is added to the test lung at rates ranging from 200-500 mL/minute to simulate a range of metabolic output. The test lung was ventilated at respiratory rates of 12-28 breaths/minute, and tidal volumes of 400-600 mL. Gas sampling is performed with a Datex-Ohmeda gas bench at the mouth of the headform after the carbon dioxide concentration in the snorkel mask reaches steady state for at least 4 minutes. The 3 anesthesia circuit filters we tested are Teleflex Main Flow Bacterial/Viral Filter 1605, ISO-GARD Filters and Filter HMEs 28012, and Romsons HME Disposable Bacterial/Viral Filter GS-2095.

In the CO2 accumulation result, for the 3 anesthesia circuit filters we tested, the steady state CO2 concentration inside the mask is approximately 1-2%, which is generally safe for short term usage [20] and comparable to commercial elastomeric respirators [22]. These preliminary results are comparable to our user feedback from University of Utah, where one of our authors self-tested and reported that the work of breathing appears similar to an N95 respirator when either filter is attached. Subjectively, it appeared comfortable but took a small adjustment period to adapt to breathing to a comfortable level. At this time, we would recommend a periodic (every 5-10 minutes) deep forced exhalation to purge the mask of any CO2 buildup, which is quite similar to previously proposed solution for elastomeric respirators [19].

As the risk of CO2 accumulation is directly related to the volume of dead-space, we performed direct volume measurement on our Pneumask-G setting with a Body Glove snorkel mask. The effective dead-space volume of the adapter is 10 mL while the effective dead-space volume of a Virex N100 In-line Filter is 11 mL, adding to a total volume of 21 mL, which is very low compared to the effective dead-space volume of the supplied snorkel (157mL, isolating inhalation/exhalation pathways). These results suggested that if a snorkel manufacturer has passed a CO2 accumulation test with their mask and snorkel tube, it is very likely that a Pneumask-G based on their snorkel mask will also pass a CO2 accumulation test.

Under the same logic, it was proposed that ventilation on relaxed or resting states may conduce to CO2 accumulation due to an insufficient quenching of the mask secondary to the low minute ventilation expected during resting states. To determine if there is a difference between resting and exercise in terms of CO2 accumulation and re-breathing, a volunteer member of the team performed a “resting test”. A male, 38 years old healthy volunteer, under standard monitoring including ECG, SatO2, NIBP, EtCO2 and Inspired CO2 measurements, wore the Pneumask in the G configuration for a total of 80 minutes. During the test, the subject lied recumbent and without moving. An anesthesiologist was present at all times during the test, recording the vital signs trend and was instructed to stop the test if vital signs deteriorated in any way. The subject could not see the monitor values to prevent from altering the normal breathing pattern in reaction to CO2 or other variable values. The results are displayed in the following spreadsheet.

Although the was an increase in the Inspired CO2, consistent with rebreathing and insufficient quenching of exhaled CO2, with a maximum value of 9 mmHg of inspired CO2 at the minute 15 of the test, the EtCO2 values remained stable, as did all the other vital signs values for the entirety of the test. Our explanation is that the inspired CO2 rise triggered an increase in minute ventilation and respiratory rate, maintaining EtCO2 within normal values. In conclusion, after 80 minutes of mask use under resting conditions, there was no significant accumulation of CO2 and no deleterious effects secondary to the observed elevation of inspired CO2. It is likely that any accumulation under resting conditions will be minimal and automatically adjusted by the user by the normal physiologic response to CO2 buildup.

### 3.5 Exercise Test

Currently, 3 exercise tests have been conducted by volunteers using the Pneumask-G configuration. In the first 2 tests, the volunteer was a 38 years old male, ASA 1 status, with weight of 83kg for a height of 1.80 m (2.03 m2 total body surface area by Mosteller formula). Both tests were performed at FiO2 of 21% (ambient inspired oxygen fraction at a barometric pressure of 1011 hPa).

In the first test, a Decathlon FreeBreath mask was used with a 1.6 cmH2O pressure drop, bacterial/viral filter with no HME (Heat and Moisture exchanger). The mask has been worn on a treadmill at maximum inclination, for 10 minutes, to measure the CO2 level during intense activity. For a constant running speed of 6mph, the inhaled CO2 remained below 2mmHg at all times, while the exhaled CO2 rises up to 48mmHg on peak physical effort.

The second test was performed by the same volunteer subject, in the same treadmill machine, under same general conditions for 1 hour, with the Pneumask-G configuration but with a HMEF filter with a pressure drop of 4.5 cmH2O, which is almost 3 times higher than the filter used for the first test. For the second test, we monitored heart rate, SpO2, non-invasive blood pressure (NIBP), ECG, End-tidal CO2, Inspiratory CO2, FiO2, as well as a number of subjective measures including discomfort and stamina. The results of this test are summarized in 22. These results indicate that the change in Inspiratory CO2 throughout use of the device, in exertion that simulates that of most healthcare work, is negligible and in line with NIOSH standards [19]. Subjective comfort/discomfort was rated from 1 of complete discomfort to 10 of complete comfort. It is notable that this never fell below a rating of a 7. Further, the volunteer had an appropriate HR response for the level of exertion and no further alterations in physiological processes were noted. This indicates the device performs similar to elastomeric respirators under near identical conditions.

The third qualitative testing was performed with Pneumask-G configuration on a Body Glove^™^ snorkel mask on one of our co-authors. A series of capnographic measurements were performed using a closed anesthesia circuit including EtCO2, pressure, volume, and flow measurements. Under normal respiration with an open mask purge valve, the EtCO2 is around 30 mmHg. Under regular simulated Operating Room activities, there was no rise in EtCO2 noted for 30 minutes. Once we closed the purge valve with tape, the EtCO2 rose to 33 mmHg with normal respiratory pattern. No rise was noted in EtCO2 with 30 minutes of regular OR activities. Work of breathing was noted to be more difficult in this setup.

### 3.6 Simulation of CO2 Species Transport and Flow Modeling (CFD)

In an effort to better understand CO2 buildup with different flow path configurations in different masks, our team has also begun to build CFD models of these masks (led by Simon Ellgas, Waymo). We are in the process of building a platform to rank different designs and mask models by theoretical relative performance.

The software used was Siemen’s STAR-CCM +; the model used the segregated flow implicit unsteady solver, with the realizable k-Epsilon URANS turbulence model. Computational runtime is around 8-9 hours per exhale-inhale cycle. We are currently solving these tests using 32 cores Intel XEON on a desktop machine (not on a compute server). Mesh size is 4.8M cells, so we have 150k cells per core. The timestep is set at 0.01s, to compromise between compute speed and quality, with a CFL number around *≈* 20.

One-way valve modeling poses some challenges numerically in the model: to avoid the numerical cost of mesh motion, and the very thin gaps present during opening and closing of the purge valves, the valves are modeled by simply varying the porous resistance of a porous region at the location of the valve. Therefore, the viscous resistance is set to a very high value to force the flow to practically zero when air would flow against the valve’s direction. For flow in the valve’s direction, the resistance is set to a value that reproduces the pressure drop across the valve in its full-open position. To be clear, the variable porous resistance is currently not set based on the local flow field at each valve, which would be more physical, but caused substantial numerical instability. Instead, each valve’s resistance parameter is set based on the global direction of the flow (inhaling, vs exhaling). This approach is only valid since the flow changes direction almost instantaneously throughout the computational domain. While this solution is pragmatic, and allows us to perform the desired qualitative ranking of the CO2 buildup of different mask configurations with quick turn-around, it can indeed be further improved.

These computational predictions of CO2, flow rates and pressures — while they appear similar in range to experiments we report here — we caution readers that these computational models were built specifically to assess relative performance of different design configurations (different flow paths, mask models, etc.) and not absolute values that could be compared to experimental tests.

We are currently extending these preliminary results to ask specific, targeted questions about relative performance of different configurations. Preliminary results comparing CO2 between the stock-snorkel mask and the Pneumask with filter and adapter are shown in Figure 23.

**Figure 23.**
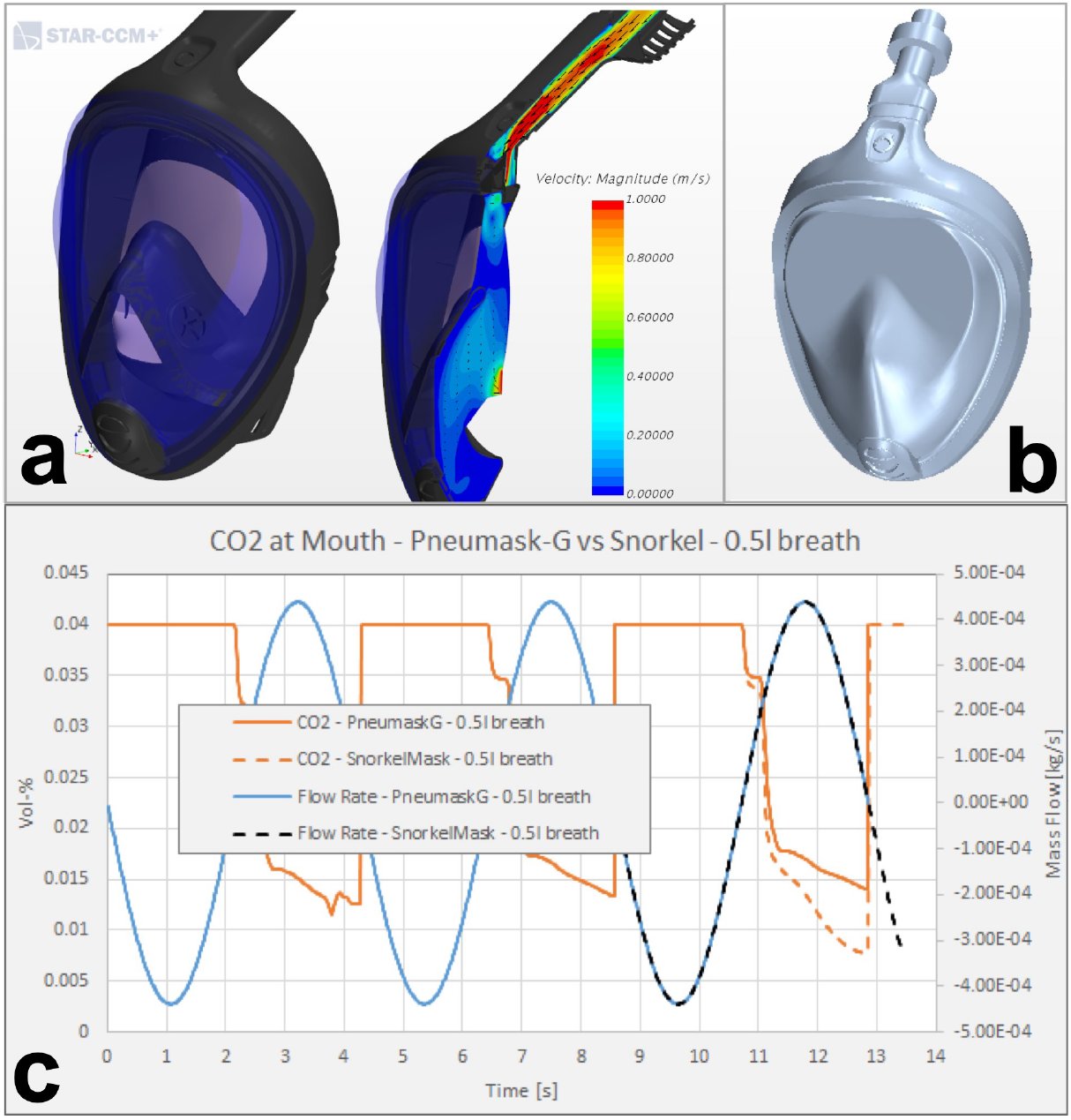
Preliminary results on flow modelling and species transport. a) Visualization of flow velocities within the Decathlon Easy Breath mask in stock configuration. b) Pneumask system with adapter and inline filter added. c) Preliminary CFD results on CO2 species transport, comparing the Pneumask with filter resistance to the stock configuration of the mask.

### 3.7 Clinical Usability Testing

The visibility of Dolfino Frontier has been extensively tested by multiple lab members, and no fogging was noted. The anesthesiologists among our co-authors also tested the same mask, and the mask did not interfere with a simulated intubation process nor with simulated standard anesthetic practices. The mask was found to reduce the volume of one’s voice when worn, which can be overcome by users increasing their volume of speaking or by using our Bluetooth microphone solution discussed further in the next section.

### 3.8 Communication Through Mask

The full-face snorkel mask can significantly muffle the user’s voice, thereby inhibiting communication and requiring the user to strain their voice to communicate with others in a noisy environment [17]. In order to help sound travel past the mask, we created a mobile app to relay audio from a Bluetooth microphone inside the mask to speakers outside the mask (Figure 24). Sound can either be played on the phone’s internal speakers or through speakers connected to the device’s wired headphone port. The download instructions and app user instructions are available in Appendix C.

**Figure 24.**
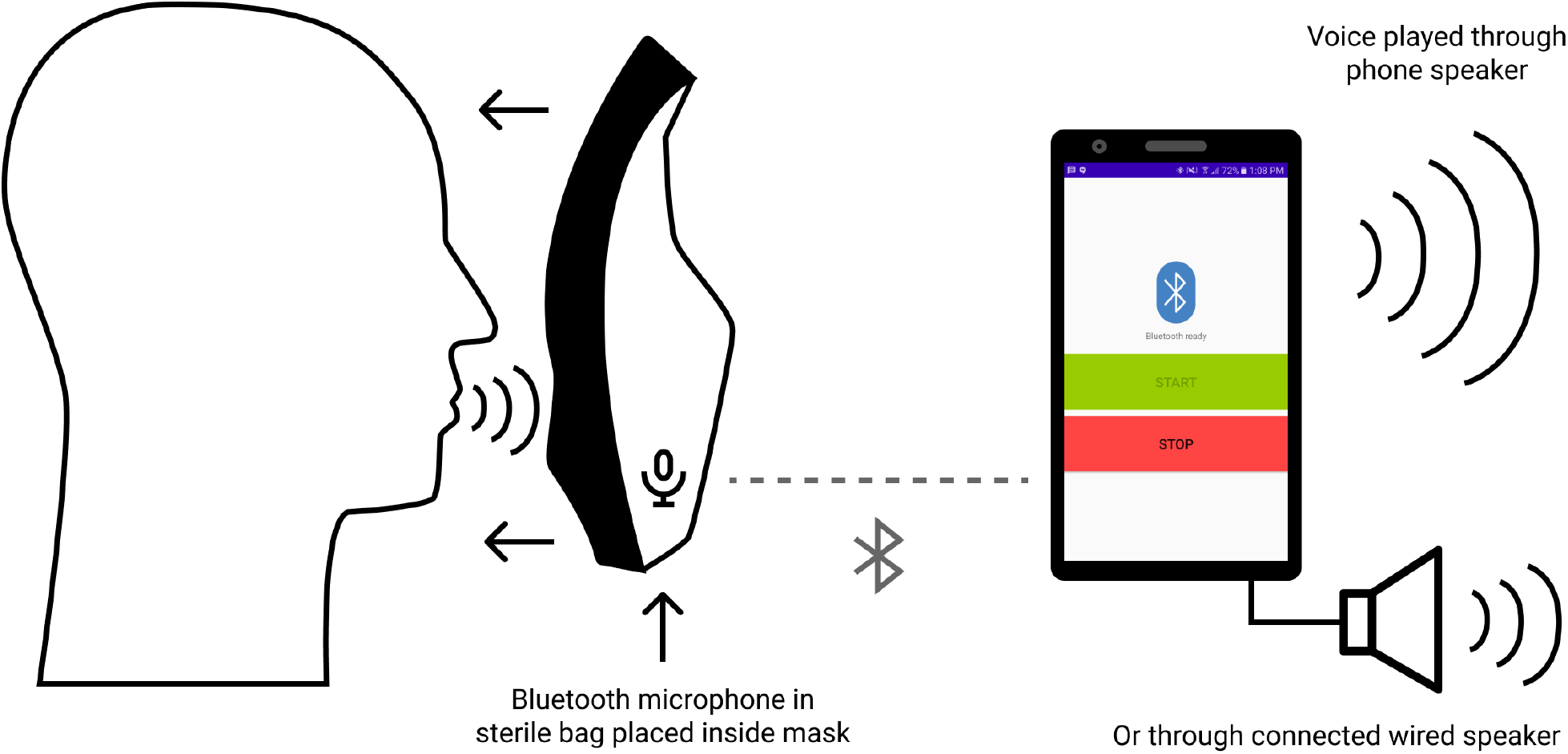
Diagram of Bluetooth amplification solution using only an android cell phone and wireless Bluetooth earbuds placed in the mouth part of the mask (can be placed in a plastic bag to make cleaning easier).

All code is available on GitHub at https://github.com/kylecombes/mic-repeater-android. This solution is currently only available on Android platforms, although the team is currently working on the iOS version, and hopes to release that soon.

## 4 Donning and Doffing Procedures

We have developed suggested donning and doffing procedures based on the recommendation of UCSF [26] and from Stanford and UCSF feedback on our prototypes. A set of suggested procedures can be found in Appendix D.

## 5 Decontamination Protocol Testing

Due to the cost and design of the full-face snorkel mask, sustainable use requires that the mask itself be reused. Thus, it will need to survive common decontamination procedures such as autoclaving or immersion in a bath of bleach or ethanol. We have performed preliminary tests in which we subjected the Dolfino Frontier mask to the conditions involved in common decontamination procedures; the mask is stated by the manufacturer to consist of either silicone or thermoplastic rubber and polycarbonate lenses.

We have developed and tested our decontamination protocols based on recommendations from the CDC [21], OSHA decontamination protocols for respirators [7], and the consensus of National Academy of Science on reusable elastomeric respirators (p. 76) [8]. From these guidelines, a simple approach could potentially be the combined usage of detergent and bleach to achieve decontamination of the snorkel masks. Besides sodium hypochlorite, there are other hospital-used disinfectants that meet the EPA’s criteria for use against SARS-CoV-2 [9] or CDC guidelines on chemical disinfectant use [10]. Among these, some hydrogen peroxide solutions, such as Accelerated Hydrogen Peroxide, offer the advantage of potentially being less harmful to the user and equipment, while only requiring a short contact time of just a few minutes. Ethylene Oxide sterilization is another commonly used method to disinfect heat sensitive equipment [11]; however, it requires specialized equipment and facilities, and whether access to such services, with the required turnaround time, is widely available to health institutions needs to be determined.

### 5.1 Autoclaving

We first performed a preliminary test to check whether mask functionality survives over the course of multiple autoclaving cycles. Before autoclaving the mask, we first took reference photos of its condition (Figure S2). We then autoclaved the mask using a 30 minute gravity cycle at 121 deg C and 15 psi, with 10 minutes of warm-up before sterilization and 30 minutes of drying afterwards. Afterwards there was a mild “hot plastic smell”. Small scratches were found upon visual inspection of the black plastic material. After letting the mask rest for at least 30 minutes to cool down, we again autoclaved the mask for another identical 30 minute gravity cycle. The mask survived both cycles of autoclaving without damage. Finally, we again let the mask rest for at least 30 minutes to cool down and then autoclaved the mask for another identical 30 minute gravity cycle. After this third round of autoclaving, with a cumulative autoclaving time of 90 minutes, the silicone rubber of the mask strap and mask seal appeared to remain elastic and functional. Mask was worn after autoclaving with no apparent loss of function.

### 5.2 Bleach Immersion

Besides autoclaving, the mask may be immersed in a bath of bleach for decontamination [12]. Thus, we tested whether a mask could survive the relatively harsh chemical conditions of immersion in a bath of bleach. We immersed a new snorkel mask for 10 hours in a bath of 10% bleach. There was no apparent damage afterwards despite some white coating which can be easily washed off (Figure S3). We thus concluded that our mask should be able to survive most bleach disinfection protocols used in the hospital [7].

### 5.3 Ethanol Immersion

Besides autoclaving and immersion in bleach, the mask may be immersed in a bath of ethanol for decontamination [12]. Thus, we tested whether a new mask could survive immersion in a bath of 70% ethanol for 10 hours (Figure S4). No apparent damage was noted afterwards. (Note that we should always use 95% ethanol to make the 70% ethanol solution, since 100% ethanol may contain trace amounts of benzene which is carcinogenic.)

### 5.4 Stretch Test

With our three snorkel masks treated under three different decontamination conditions (3 cycles of autoclaving, 10 hours of bleach immersion, and 10 hours of ethanol immersion, respectively), we then performed a simple stretch test on the elastomer bands of each mask by holding the strap with both hands such that the thumbs touched each other at the tips, then pulling and qualitatively observing the separation. The straps for the ethanol-treated mask appeared to have stretched the most, while the straps for the bleach-treated mask appeared to have stretched the least. Nonetheless, all the masks are functional and seal well after all the cleaning processes.

### 5.5 Dry Heat at 65C

Several reports indicate that dry heat at 65 degrees C is capable of killing any viral particles [13, 14]. Although we did not explicitly test this protocol, given the fact that the masks survived 121 degrees C in the autoclave for 30 minutes, we can safely infer that our mask will also survive a dry heat disinfection protocol.

### 5.6 Decontamination Summary - Guidelines

Based on above testing results and the recommendation from OSHA [7], we developed suggested protocols for cleaning and decontaminating our snorkel mask (Dolfino Frontier), which is available in Appendix B. Please note that this protocol is not formally approved, and each hospital should consult their EH&S officers or infection disease specialists for a standard operating procedure. If you are using disinfectants other than bleach, please also check this table compiled by EPA for recommended cleaning time.

Another important note is that not all snorkel masks can tolerate the decontamination. For example, snorkel masks from Animdive, Tinmiu and Keystand cannot withstand the temperature of autoclave or industrial washers commonly used in OR. If you are using snorkel masks other than Dolfino Frontier, please perform appropriate testing before usage.

## 6 Failure Modes and Effects Analysis (FMEA)

We have also performed a failure modes and effects analysis on our Pneumask-G design. In this analysis, we first decomposed the product into different components and listed the primary functions of each component. We then analyzed what would happen if each component fail to serve their primary functions and how much negative impact it would bring. By considering the severity, chance of occurrence, and chance of detection of that malfunctioning scenario, we can compute a semi-quantitative score. By comparing the semi-quantitative score of each possible failure mode, we can identify the most important failure modes that require immediate action or improvement in design.

The analysis suggested several points that may be useful for anyone to further build upon our system: (1) Using surgical hood or any additional coverage to protect the filter surface, lateral side of the mask and the straps from gross contamination may provide additional protection (2) Separating the airflow pathway for inhaled air and exhaled air as much as possible can further improve performance for the Pneumask-G design. Modification or blocking of the chin valve would require complete redesign of airflow pathway. (3) Use a single-usage filter if available. For filters designed for repetitive usage, follow the instructions and regulatory-approved extended use claims (EUC) of the manufacturers (4) Avoid prolonged usage if possible (5) Long-term durability of the adapter when subjected to many cleaning process cycles will likely be dependent on the exact material and manufacturing process. Please perform appropriate further failure testing to characterize usage lifetime of these adapters in the material that is used. (6) Instructions on cleaning should specifically mention the necessity of cleaning the valve, as discussed in step 5 of the decontamination protocol in Appendix B. (7) Use of voice amplifying system may help to minimize risk due to jaw movements (8) Encourage face washing after doffing.

## 7 Conclusions

We recognize that under normal circumstances, the adaptation of recreational sports equipment for medical usage would not be advisable, because of the availability of superior alternatives (and more medically-specific in their overall design). A recent survey [3] shows that in the US, 31.4% of health care workers reported that there are no masks in their hospitals. The number of healthcare workers without access to suitable PPE will continue to grow if no additional efforts are made, due to a discrepancy between demand and available supply. With lack of PPE solutions for health care workers globally, new alternatives have to be explored. **But even more critical than the design of these new solutions, is the rigorous and stringent evaluation of quantitative performance**. Here we perform stringent testing on topics ranging from CO2 accumulation, fit testing, filtration efficiency testing and valve performance.

- Our tests found the Pneumask capable of forming a seal that exceeds the standards required for half-face respirators or N95 respirators, through both quantitative and qualitative fit testing. We found this to be the case for both the Dolfino Frontier and the Subea Decathlon mask models. We encourage readers who wish to replicate our fit-testing results to perform quantitative fit testing with a very high performance HEPA-rated inline filter (standard is 99.97% efficient for particles at 0.3 *μ*m). This is to ensure that the fit of the mask (*sealing capability of the mask*) is evaluated separately from the filter performance. If no filter is available that operates >99% efficiency at breathing flow-rates, you may need to use specialized modes for fit testing (such as N95 mode on Portacount devices).
- Work of breath on the overall system was evaluated qualitatively to be comfortable to the user with several different filter types for extended periods (1-3 hours).
- Filter testing indicates a range of options with varying performance depending on the quality of filter selected, but with typical filter performance exceeding or comparable to the N95 standard. If multiple filter options are available to clinicians, we recommend the usage of inline pleated, hydrophobic mechanical filters (such as the Pall BB25 or BB50T) rather than electrostatic/viral filters. This is due to superior stated filtration performance in the specification of the filter, and also due to superior filter durability. Extended use of filters in this context is still under evaluation. See table 15.
- CO2 buildup was found to be roughly equivalent to levels found in half-face elastomeric respirators in literature; between 1 and 2 percent.
- We report results for average exhalation valve leakage of 1.5 mL/min at a suction of 25mm water-column height over 12 tests. This result indicates good performance when compared to the maximum NIOSH specification exhale valves (30 mL/min) at this pressure. We report a theoretical estimation of valve closure time of approximately 0.024 seconds.
- Clinical usability tests indicate sufficient visibility. Clinical usability tests indicate speaking can be muffled, especially in noisy environments. In these environments, this muffled speech poses a technical challenge for users. We have developed an open-source amplification solution using hardware that many clinicians own personally: a Bluetooth headset (or earbud) and a smartphone. This solution is optional for users, and will depend on individual preferences and occupational circumstances. The android app is available, the iOS version is still under development.
- While not intended for extreme extended usage, durations on the order of 4-6 hours of continuous use in clinical environments have been reported by international partners (in France and Chile).
- We present guidance on the assembly, usage (donning and doffing) and decontamination protocols. Testing of decontamination protocols indicate reuse of the snorkel masks is technically feasible.

Additionally, we recognize that beyond being an alternative to N95 respirators (and protective goggles/face shields), Pneumasks can be combined with a disposable hood to leave no directly exposed hairs or skins that are otherwise susceptible to being contaminated with droplets (Figure 25). Without Pneumasks, this kind of protection is only achievable with PAPR or the use of surgical hood/coveralls, which may also be in short supply and are associated with a more complex doffing process [27].

**Figure 25.**
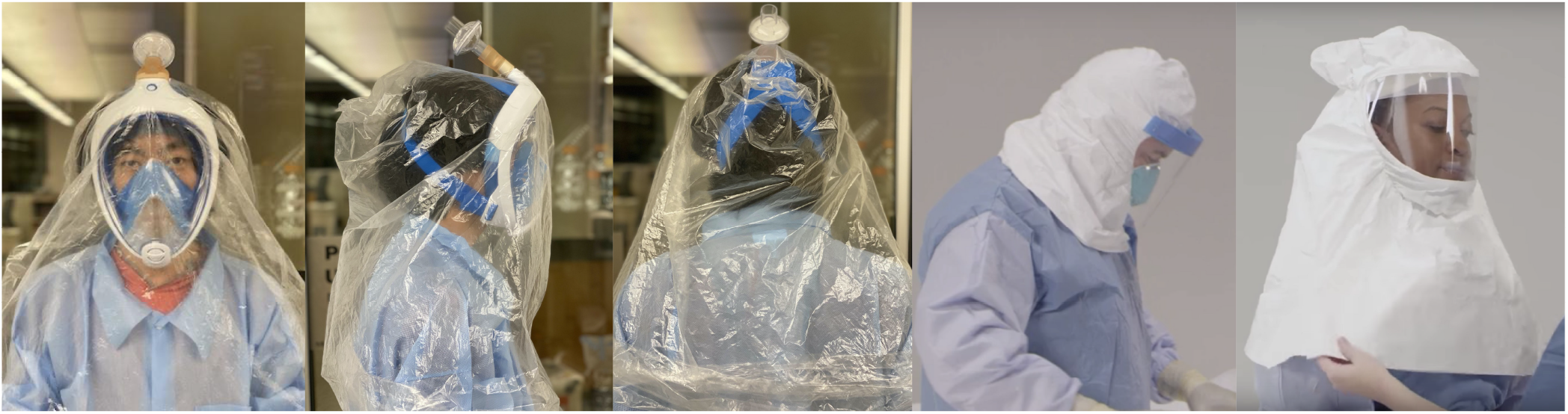
Pneumasks can be used with a disposable plastic hood to offer additional droplet protection. Also shown as references are uses of surgical hood and PAPR [27].

**Figure 26.**
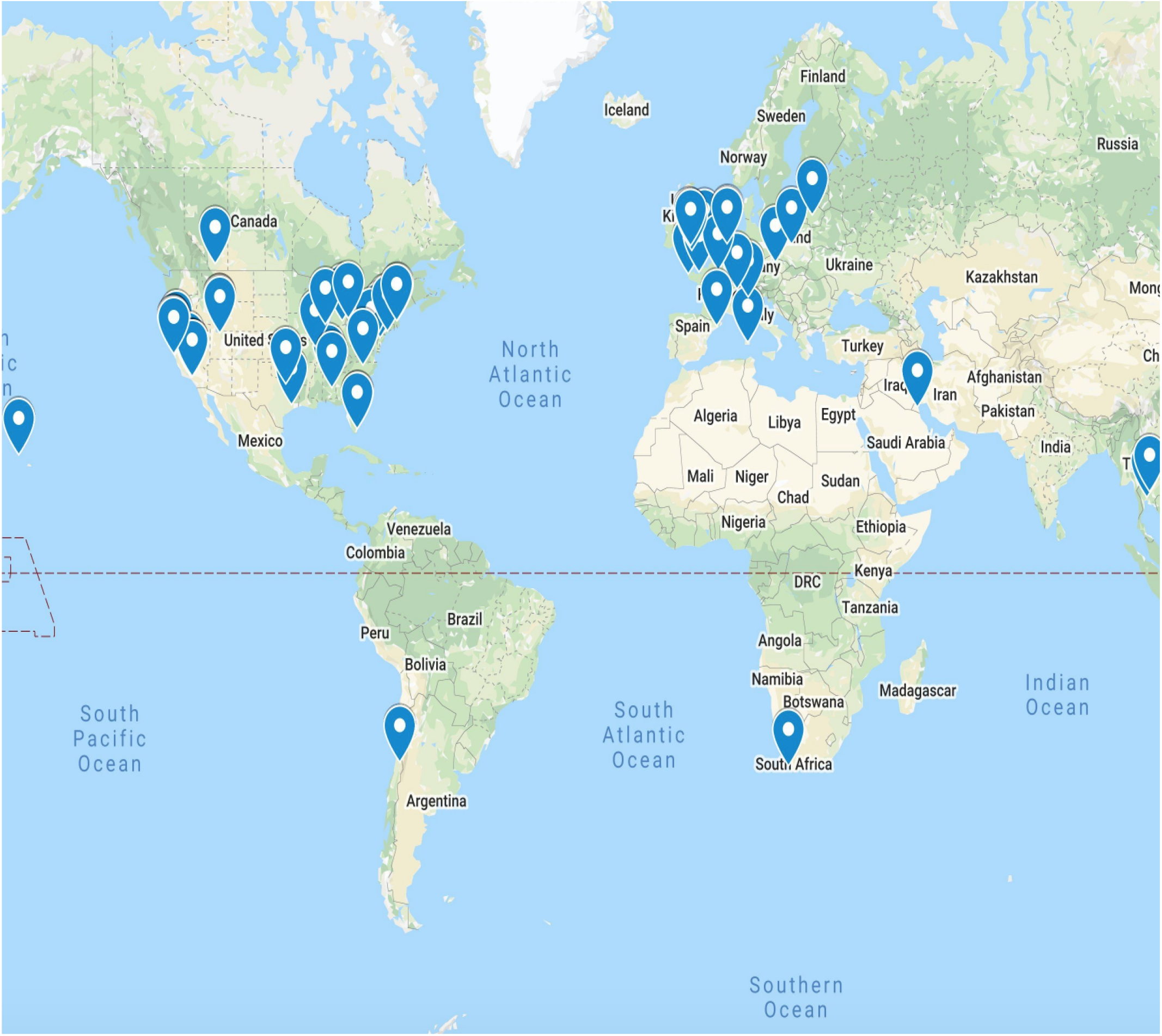
Map showing where our collaborators are throughout the world.

**Figure 27.**
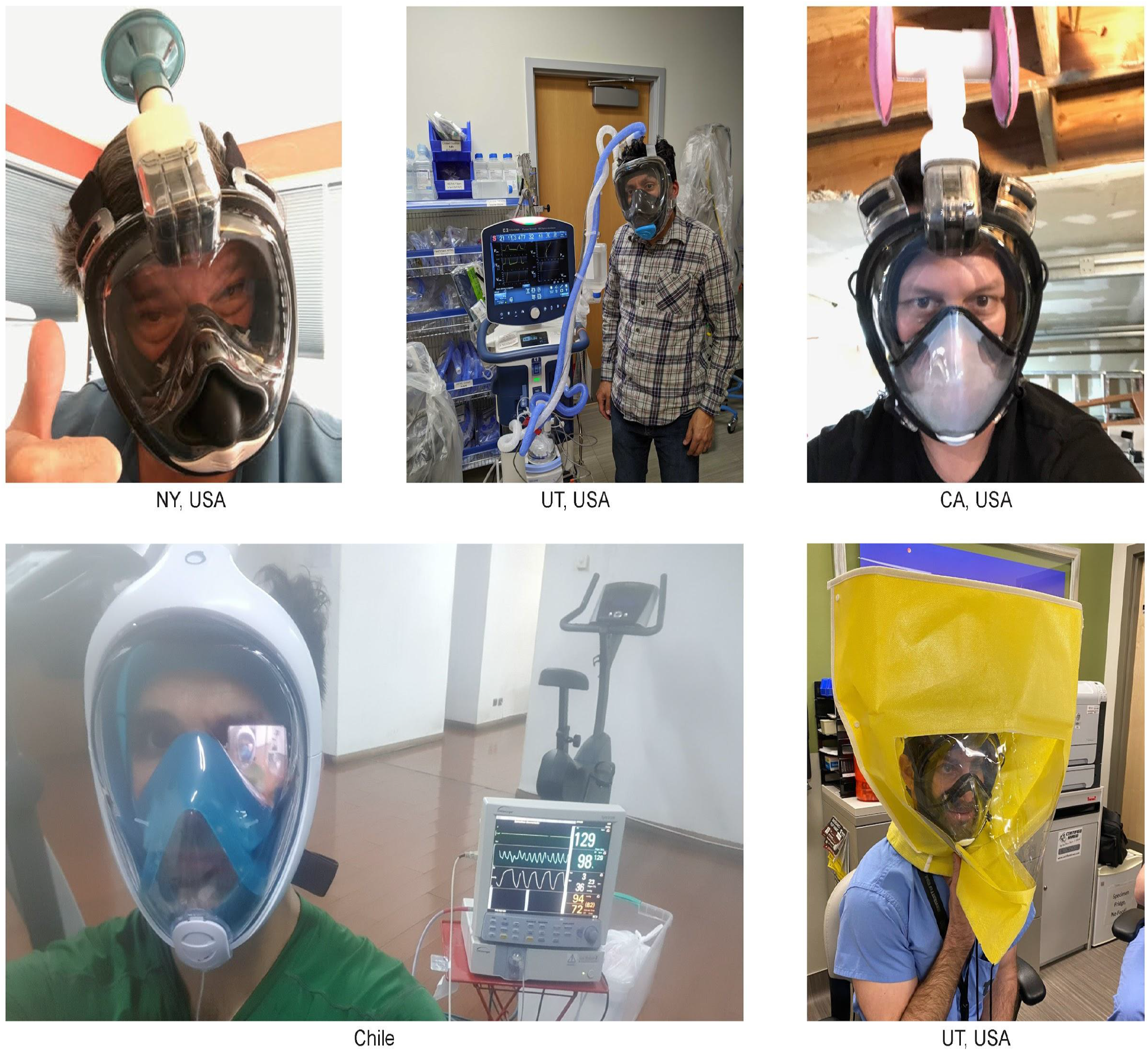
Global efforts on design effort by our team of collaborators worldwide.

**In the context of a dire worldwide shortage of PPE for medical personnel, and where no other approved alternatives are available, we are cautiously optimistic about the performance and efficacy of this system as an N95-alternative technology**.

## 8 Regulatory Guidance

The FDA has issued Emergency Use Authorization (EUA) for PPE, including NIOSH approved respirators, which has significantly altered the clearance process for solutions such as ours. We are actively seeking guidance from the FDA for our approach in order to provide regulatory clarity to our partners and clinical sites to expedite implementation. It is our understanding that this approach will allow for NIOSH approved filtration and meet or exceed fit specifications for N95 and elastomeric respirators currently authorized by the EUA.

Upon further discussions with the FDA, we have received the following communication regarding use solutions such as the Full-Face Snorkel Mask PPE:

FDA recognizes the urgent need for face masks in the setting of the COVID-19 pandemic due to increased use and shortages in their availability.

FDA does not object to the marketing and distribution of face masks in the healthcare setting without prior 510(k) clearance if the product is labeled in the following manner:

1. It states it may be used when FDA cleared masks are unavailable;
2. It recommends against use in a surgical setting or where significant exposure to liquid bodily or other hazardous fluids may be expected;
3. It makes no claims of antimicrobial or antiviral protection;
4. It makes no claims of infection prevention or reduction;
5. It makes no claims regarding flammability
6. The labeling contains a list of the body contacting materials.
7. The mask is not labeled as a “surgical mask”; rather it may be labeled as a “face mask”

In addition, FDA does not intend to object to marketing of masks that meet the above criteria even if they are manufactured at facilities that do not meet 21 CFR 820.

As an update, on 3/25/2020, the US FDA issued updated guidance titled “Enforcement Policy for Face Masks and Respirators During the Coronavirus Disease (COVID-19) Public Health Emergency”(FDA-2020-D-1138). In summary, this guidance now covers solutions such as the Reusable Full-Face Snorkel Mask PPE project with the following (pg3):

> FDA recognizes that, when alternatives, such as FDA-cleared masks or respirators, are unavailable, individuals, including healthcare professionals, might improvise PPE. FDA does not intend to object to individuals’ distribution and use of improvised PPE when no alternatives, such as FDA-cleared masks or respirators, are available.

The CDC has published guidelines on Crisis/Alternate Strategies for PPE decisions in the setting of N95 respirator shortages. At this time, our solution can be considered a case of “HCP use of non-NIOSH approved masks or homemade masks” which is permissible in a setting where N95 respirators are so limited that routine use of them is no longer possible and surgical masks are not available. While we believe and have documented that our solution can provide higher levels of protection than homemade or simple cloth masks, we cannot make legal claims at this time until further guidance is received from regulatory authorities.

## 9 Collaborative Efforts

This project was a global collaborative effort that crossed typical boundaries between academic, industry, government and medical institutions. Convergent design on this concept occurred nearly simultaneously in many different regions throughout the world on this work, and it brought together a highly diverse and driven community. For more information on the consortium of industrial and institutional contributors to this project who are involved with the development and distribution of Pneumask in the United States, please reference: www.pneumask.org. For Europe and France, see involved entities here: (https://adaptateur-masque.planktonplanet.org/).

## Data Availability

All data is available in the manuscript and upon request from the authors.

## 10 Acknowledgements

This has been a large community projects with support from global community including clinicians who have given us feedback throughout. We thank everyone who has engaged with the Pneumask community, specially international community working to build solutions for the current PPE shortage. We thank Schmidt Futures, Moore Foundation, Autodesk and CZ BioHub for financial support of the project. We thank Boston Scientific and Medtronics staff for support in development of injection molding parts and overall consultation. We thank MITRE and Helena for conversations around equitable distribution and members from Pall corporation for discussions around filter efficiency.

## 11 Conflict of interest

Gerry Ayala (Wildhorn Outfitters), Quentin Allinne (Subea Decathlon), and Dave Kasper (iSnorkel Inc) have company affiliations with snorkel-mask manufacturers or distributors. They consulted on original designs of these masks, and in some cases to help with adapter design strategy. None of the scientific data included in this report, nor the conclusions of this report were written or substantially influenced by these co-authors.

John Pearson, MD has a conflict of interest, as a stake holder in a for-profit entity focused on snorkel-based PPE. Patrick Kolbay is volunteering his time at this company.

We certify that none of the other authors have any affiliations with or involvement in any organization or entity with any financial interest in the subject matter or materials discussed in this manuscript. None of the other authors (beyond what is listed above) have any affiliations with John Pearson’s company.

## 12 Publisher’s note

This is an early preprint of this article intended for an preprint online platform. This article is intended for submission to a peer-reviewed journal at a later date. We are sharing this openly to enable others to find and replicate this work and engage to further improve this in an open manner.

## 14 Appendices and Supplementary information

Please reference the below link for hyperlinked tables (at the end of this live Lab-notebook): https://docs.google.com/document/d/1J22le3dBZBnNDXGlJLRb38z7v7LaOjKfDeN9f0tFeKY/edit?usp=sharing, but are also shown below.

## 14.1 Appendix A Supplementary Tables

**Table S1, Part 1 of 2:**
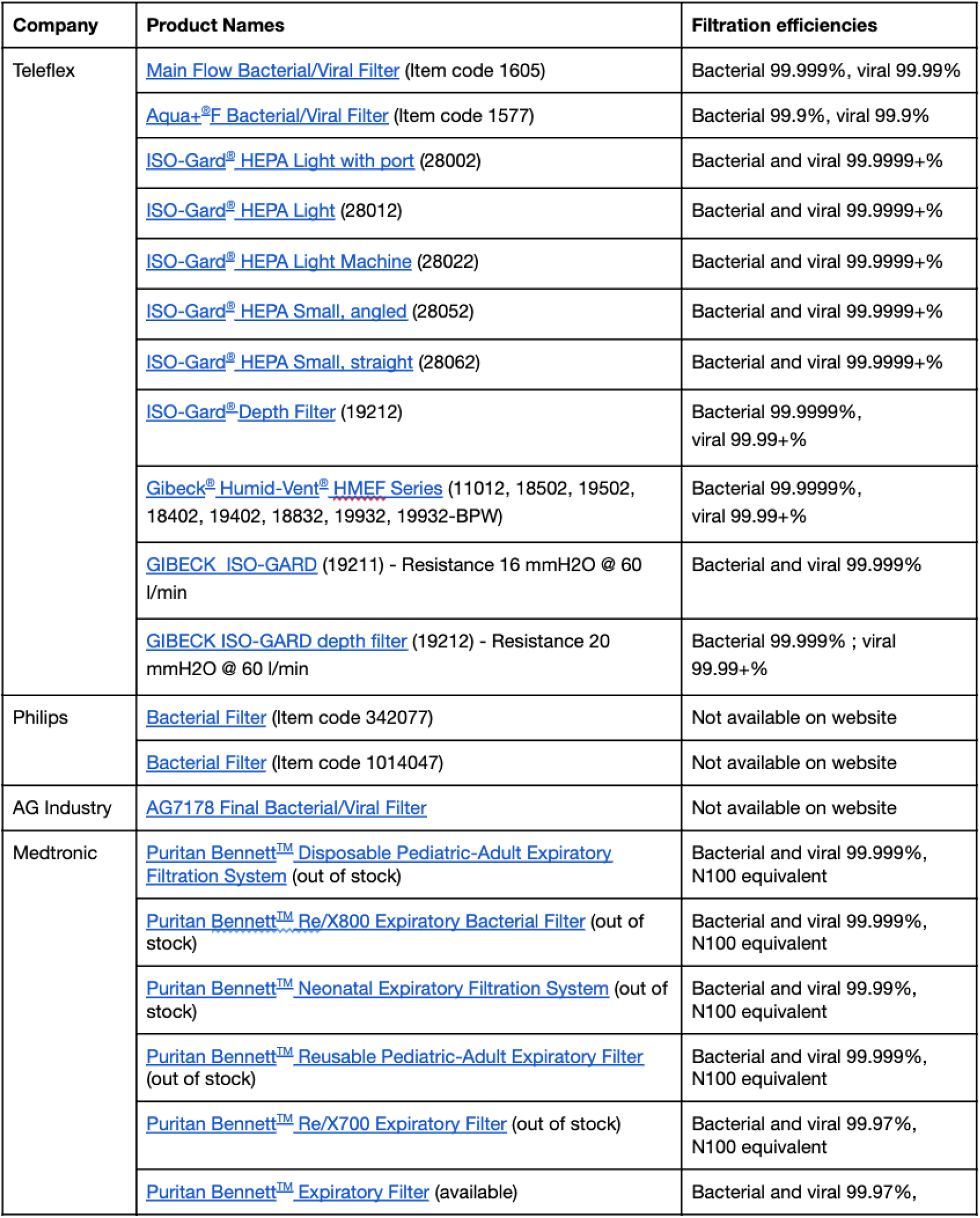
List of commercially available respiratory filters which are initially designed for ventilators.

**Table S1, Part 2 of 2:** List of commercially available respiratory filters which are initially designed for ventilators.

Table S1, Part 1 of 2: List of commercially available respiratory filters which are initially designed for ventilators.
|  |  |  |
| --- | --- | --- |
|  |  | N100 equivalent |
|  | Puritan Bennett™ D/X800 Expiratory Filter (out of stock) | Bacterial and viral 99.999%, N100 equivalent |
| Airon | Bacterial/Viral Filter (P/N 58210) | Not available on website |
| PALL Corporation | Pall BB50T Breathing circuit filter (Item code BB50T) | Bacterial and viral 99.999% |
|  | Ultipor® 25 Filter With Monitoring Port (Item code BB25AB) | Bacterial and viral 99.999% |
| GE Healthcare<br>(filters not available now as confirmed in email) | HMEF Neonatal bacterial viral HMEF + Luer Port (2106570-006) | Bacterial and viral >99.9% |
|  | Pediatric bacterial viral HMEF + Luer port (2106570-009) | Bacterial and viral 99.99% |
|  | Adult bacterial viral HMEF + Luer port (2106570-010) | Bacterial and viral 99.999% |
|  | Adult HEPA pleated paper hydrophobic bacterial viral filter + Luer port (2106570-008) | Bacterial and viral 99.99999% |
|  | Adult bacterial viral filter + Luer port (2106570-007) | Bacterial and viral 99.999% |
| DAR | Mechanical Filter (Small) | Bacterial and viral 99.999% |
| GVS Filter Technologies | Electrostatic Filter without gas sampling port | Bacterial and viral 99.999% |
|  | Mechanical HEPA filter Long Term anesthesia-ventilation | Bacterial 99.999% - viral 99.997% |
|  | Reusable Hepa Filter - Autoclavable up to 20 times | Bacterial and viral 99.999% |
|  | Reusable Hepa Filter | Bacterial and viral 99.999% |
|  | Multi Vent Barb - Multi-ventilator filter | Bacterial and viral 99.999% |
|  | Electrostatic Filter 22M-15F-22F | Bacterial and viral 99.999% |
|  | Bacterial-Viral filter for nebulizer therapy machine | Bacterial and viral 99.999% |
|  | Filter for nebulizer therapy machine | Bacterial and viral 99.999% |
|  | Maxi Pleated HEPA Filter - Resistance 142 Pa | Bacterial and viral 99.9999% |
| Vyaire Medical | Hygroscopic Condenser Humidifier with Filter AirLife® Vyaire Medical 5708 | Bacterial: 99.997%<br>Viral: 99.94% |

**Table S2, Part 1 of 1:**
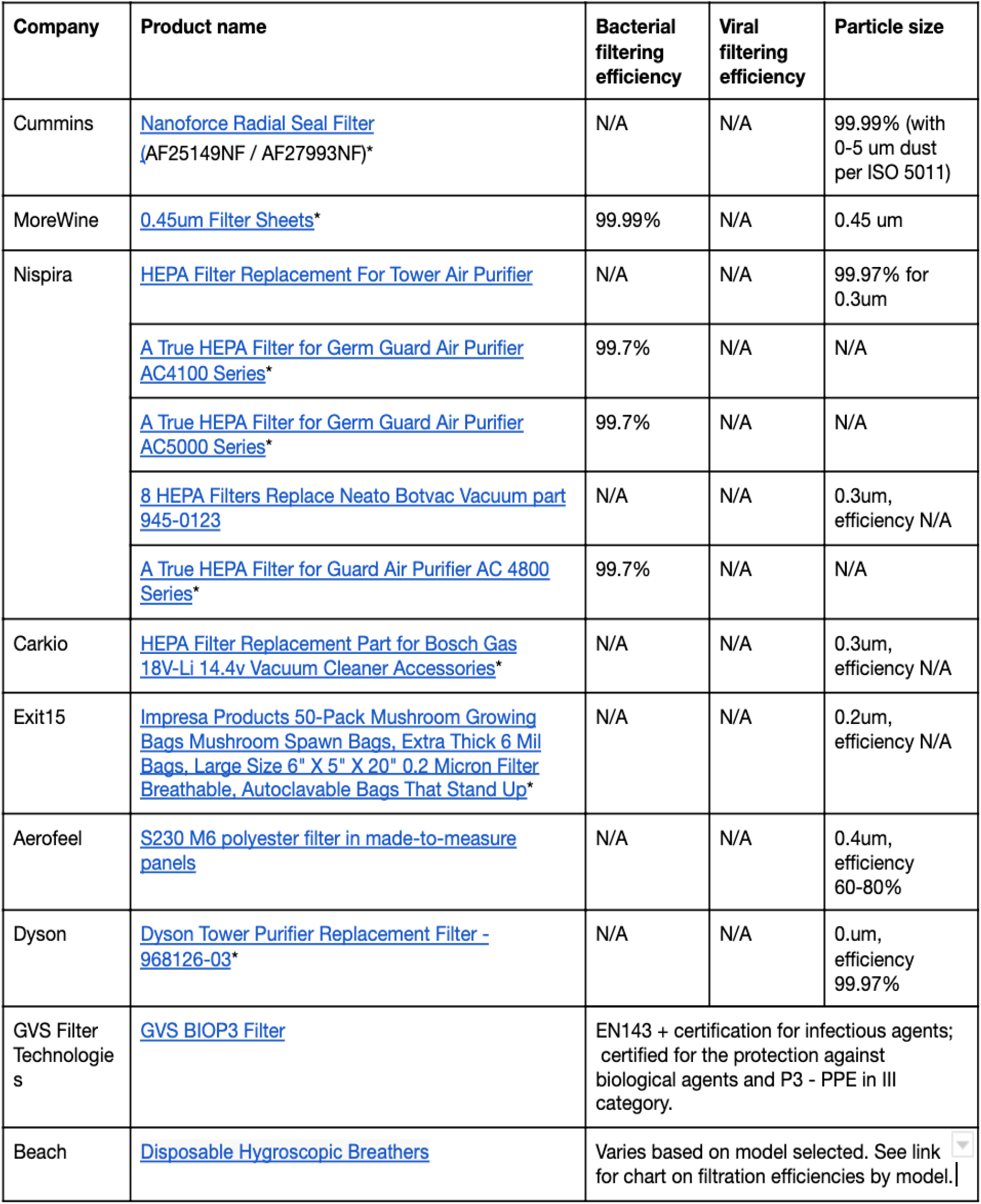
List of commercially available filters which are not initially designed for medical usage. More characterizations of the filtering abilities are required before using in a hospital setting. The star marks next to the product name indicate the ones Prakash Lab is planning to test. (N/A = not available.)

Table S1, Part 2 of 2: List of commercially available respiratory filters which are initially designed for ventilators.
| Company | Product name | Bacterial filtering efficiency | Viral filtering efficiency | Particle size |
| --- | --- | --- | --- | --- |
| Cummins | Nanoforce Radial Seal Filter (AF25149NF / AF27993NF)* | N/A | N/A | 99.99% (with 0-5 um dust per ISO 5011) |
| MoreWine | 0.45um Filter Sheets* | 99.99% | N/A | 0.45 um |
| Nispira | HEPA Filter Replacement For Tower Air Purifier | N/A | N/A | 99.97% for 0.3um |
|  | A True HEPA Filter for Germ Guard Air Purifier AC4100 Series* | 99.7% | N/A | N/A |
|  | A True HEPA Filter for Germ Guard Air Purifier AC5000 Series* | 99.7% | N/A | N/A |
|  | 8 HEPA Filters Replace Neato Botvac Vacuum part 945-0123 | N/A | N/A | 0.3um, efficiency N/A |
|  | A True HEPA Filter for Guard Air Purifier AC 4800 Series* | 99.7% | N/A | N/A |
| Carkio | HEPA Filter Replacement Part for Bosch Gas 18V-Li 14.4v Vacuum Cleaner Accessories* | N/A | N/A | 0.3um, efficiency N/A |
| Exit15 | Impresa Products 50-Pack Mushroom Growing Bags Mushroom Spawn Bags, Extra Thick 6 Mil Bags, Large Size 6" X 5" X 20" 0.2 Micron Filter Breathable, Autoclavable Bags That Stand Up* | N/A | N/A | 0.2um, efficiency N/A |
| Aerofeel | S230 M6 polyester filter in made-to-measure panels | N/A | N/A | 0.4um, efficiency 60-80% |
| Dyson | Dyson Tower Purifier Replacement Filter - 968126-03* | N/A | N/A | 0.um, efficiency 99.97% |
| GVS Filter Technologies | GVS BIOP3 Filter | EN143 + certification for infectious agents; certified for the protection against biological agents and P3 - PPE in III category. |  |  |
| Beach | Disposable Hygroscopic Breathers | Varies based on model selected. See link for chart on filtration efficiencies by model. \| |  |  |

**Table S3, Part 1 of 1:** List of commercially available full-face snorkel masks.

| **Brand** | **Model (with link)** | **Size range** | **Price (retail)** |
| --- | --- | --- | --- |
| Cressi | Duke Dry | S/M, M/L | 79.95 |
| Easy Snorkel | PRODIGY | S/M, L/XL | 35.99 |
| Greatever | G2 | S, L | 36.99 |
| Guardian | Ocean See | S/M | 69.95 |
| Head | Sea Vision | S/M, L/XL | 64 |
| Head by Mares | Seaview | XS/S, S/M, L/XL | 54.99 |
| HKJB | Full Face K2 | XS, S, L | 39.98 |
| Jogoo | (180° View Snorkel Mask) | S/M, L/XL | 19.99 |
| Meduzi | Seafin | ? | ? |
| Midry | Midry | ? | 39.97 |
| Ocean Reef | Aria Classic | S/M, M/L | 69.95 |
| Ocean Reef | Aria Junior | n/a | 49.95 |
| Ocean Reef | Aria QR+ | S/M, M/L, L/XL | 89.95 |
| Ocean Reef | Uno | S/M, L/XL | 60 |
| OctoberMoon | (180° snorkel mask) | ? | ? |
| Outdoor Master (dba Greatover) | (180° snorkel mask) | ? | ? |
| QingSong | (Full face snorkel mask) | S/M, L/XL | 35.99 |
| Seabeast | AF90 | S/M, L/XL | 59.99 |
| Seac USA | Magica | Junior, XS/S, S/M, L/XL | 46 |
| Seac USA | Unica | Junior, XS/S, S/M, L/XL | 69 |
| Seac USA | Libera | Junior, XS/S, S/M, L/XL | 79 |
| Swordfish Sport | Fiji | S/M, L/XL | 76 |
| Tribord (brand owned by Decathlon S.A.) | Easybreath | XS, S/M, M/L | 89.9 |
| Trimagic | (180° Full face Snorkel mask) | XS, S, M, L, XL | 24.99 |
| U.S. Divers | Airgo LX | S, M, L | 69.99 |
| Vaincre | (180° Full Face Snorkel Mask) | S/M, L/XL | 24.99 |
| Wet Products | (Adult Full Face Snorkel Mask) | n/a | 59 |
| WildHorn Outfitters | Seaview 180° V2 | XS, S/M & M/L | 79.99 |
| WildHorn Outfitters | Seaview 180° | S/M, L/XL |  |
| X-Lounger | (Snorkel Mask Full Face Foldable) | M, L | 27.96 |

**Table S4, Part 1 of 2:** List of commercially available P100 respirator filters/cartridges originally for industrial usage.

**Table S3, Part 1 of 1: List of commercially available full-face snorkel masks**
| Company | Product name | Special Notes |
| --- | --- | --- |
| 3M | 3M™ Particulate Filter 2091/07000(AAD), P100 100 EA/Case | User manual |
|  | 3M™ Particulate Filter 2096, P100, with Nuisance Level Acid Gas Relief 100 EA/Case |  |
|  | 3M™ Particulate Filter 2097/07184(AAD), P100, with Nuisance Level Organic Vapor Relief 100 EA/Case | User manual |
|  | 3M™ Advanced Particulate Filter 2291, P100 100 EA/Case |  |
|  | 3M™ Advanced Particulate Filter 2296, P100, with Nuisance Level Acid Gas Relief 100 EA/Case |  |
|  | 3M™ Advanced Particulate Filter 2297, P100, with Nuisance Level Organic Vapor Relief, 100 EA/Case |  |
|  | 3M™ Particulate Filter 7093, P100 60 EA/Case |  |
|  | 3M™ Particulate Filter 7093B, P100 144 EA/Case |  |
|  | 3M™ Hydrogen Fluoride Cartridge/Filter 7093C, P100, with Nuisance Level Organic Vapor and Acid Gas Relief, 60 EA/Case |  |
|  | 3M™ Organic Vapor Cartridge/Filter 60921, P100 60 EA/Case |  |
|  | 3M™ Organic Vapor/P100 Service Life Indicator Cartridge 60921i |  |
|  | 3M™ Acid Gas Cartridge/Filter 60922, P100 60 EA/Case |  |
|  | 3M™ Organic Vapor/Acid Gas Cartridge/Filter 60923, P100, 60 EA/Case |  |
|  | 3M™ Ammonia/ Methylamine/Filter 60924, P100, 60 EA/Case |  |
|  | 3M™ Formaldehyde Organic Vapor Cartridge/Filter 60925, P100 60 EA/Case |  |
|  | 3M™ Multi Gas/Vapor Cartridge/Filter 60926, P100 60 EA/Case |  |
|  | 3M™ Organic Vapor/Acid Gas Cartridge/Filter 60928, P100 60 EA/Case |  |
|  | 3M™ Mercury Vapor/Chlorine Gas Cartridge/Filter 60929S, P100, 60 EA/Case |  |
| MSA | Flexi-Filter P100 818342 |  |
|  | Flexi-Filter P100 with nuisance level AG, HF Removal 818344 |  |
|  | Flexi-Filter N95 818346 |  |
|  | Flexi-Filter N95 with Nuisance Level OV Removal 818347 |  |
|  | Advantage Chemical and Combination Cartridges P100 10123079 |  |
|  | Cartridges, ADV, GMA-P100 815362 |  |
|  | Cartridges, ADV, GMB-P100 815363 |  |
|  | Cartridges, ADV, GMC-P100 815364 |  |

**Table S4, Part 2 of 2:** List of commercially available P100 respirator filters/cartridges originally for industrial usage.

|  |  |  |
| --- | --- | --- |
|  | Cartridges, ADV, GMD-P100 815365 |  |
|  | Advantage Cartridges, Multigas/P100(GME) 815366 |  |
|  | Cartridges, ADV, MERSORB-P100 815368 |  |
|  | Cartridges, Advantage, GMI-P100 815641 |  |
|  | Comfo Respirator Cartridge, Low-Profile P100 10153412 |  |
|  | Cartridges, Comfo, Sprkfoe, P100 815176 |  |
|  | Cartridges, Comfo L-profile, P100 815177 |  |
|  | Cartridges, Comfo, GMA-P100 815178 |  |
|  | Cartridges, Comfo, GMB-P100 815179 |  |
|  | Cartridges, Comfo, GMC-P100 815180 |  |
|  | Cartridges, Comfo, GMD-P100, 815181 |  |
|  | Cartridges, Comfo, GME-P100 815182 |  |
|  | Cartridges, GMI-P100 815184 |  |
|  | Cartridges, Comfo, Mersorb-P100 815185 |  |
|  | Cartridges, Comfo, GMA-P100 815186 |  |
|  | Cartridges, Comfo, GMC-P100 815188 |  |
| Honeywell | Honeywell P100 Filter Replacement Kit, For Honeywell Convenience Pack Respirators, RWS-54042 | Product guide |

**Table S5, Part 1 of 2:**
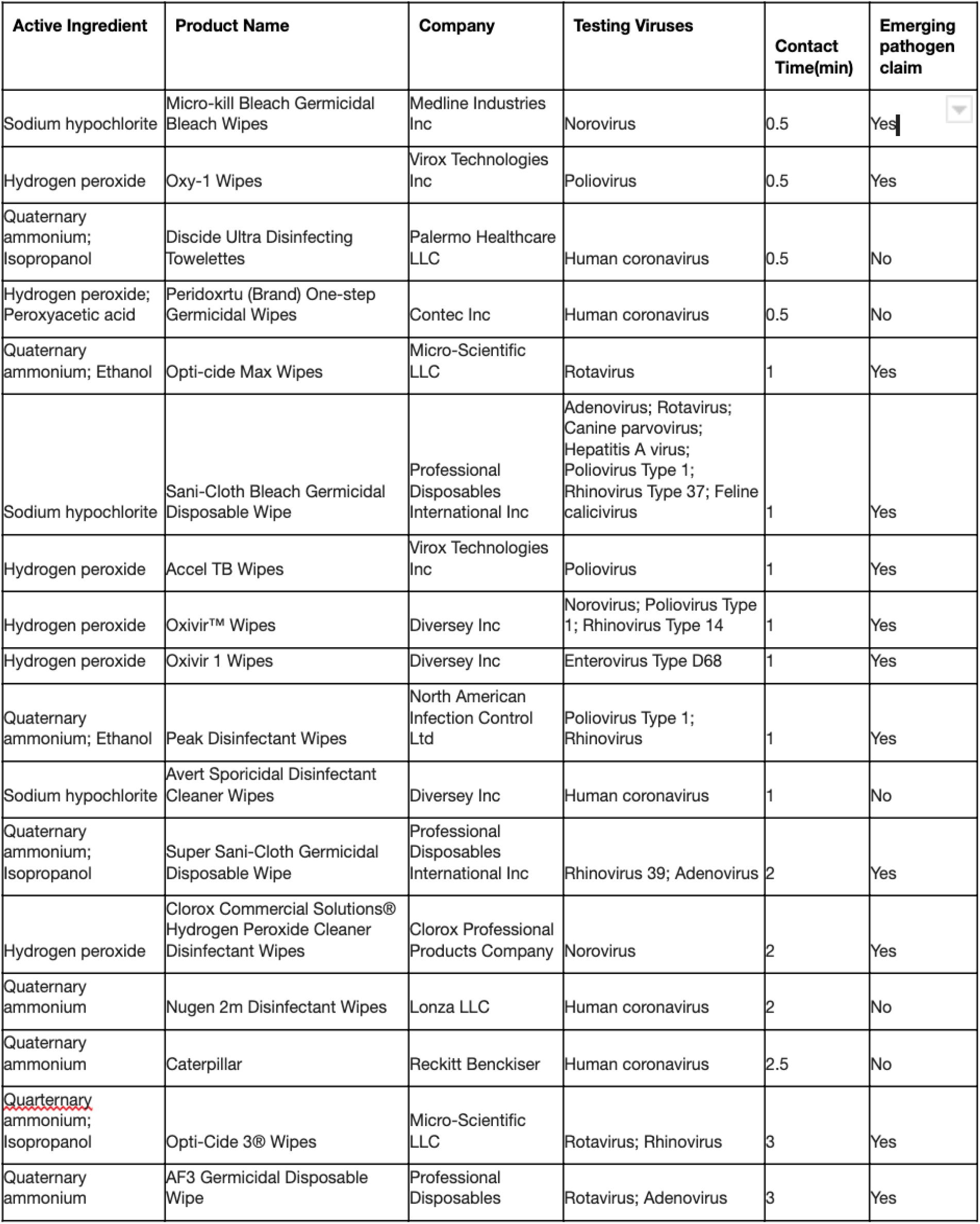
List of EPA-certified wipes. It is ordered by required contact time. All listed products are approved for healthcare use. If a product qualifies for the emerging viral pathogen claim, it is effective against a harder-to-kill virus than human coronavirus. All products on this list meet EPA’s criteria for use against SARS-CoV-2, including those marked as “No” in this column.

| Active Ingredient | Product Name | Company | Testing Viruses | Contact Time(min) | Emerging pathogen claim |
| --- | --- | --- | --- | --- | --- |
| Sodium hypochlorite | Micro-kill Bleach Germicidal Bleach Wipes | Medline Industries Inc | Norovirus | 0.5 | Yes |
| Hydrogen peroxide | Oxy-1 Wipes | Virox Technologies Inc | Poliovirus | 0.5 | Yes |
| Quaternary ammonium; Isopropanol | Discide Ultra Disinfecting Towelettes | Palermo Healthcare LLC | Human coronavirus | 0.5 | No |
| Hydrogen peroxide; Peroxyacetic acid | Peridoxrtu (Brand) One-step Germicidal Wipes | Contec Inc | Human coronavirus | 0.5 | No |
| Quaternary ammonium; Ethanol | Opti-cide Max Wipes | Micro-Scientific LLC | Rotavirus | 1 | Yes |
| Sodium hypochlorite | Sani-Cloth Bleach Germicidal Disposable Wipe | Professional Disposables International Inc | Adenovirus; Rotavirus; Canine parvovirus; Hepatitis A virus; Poliovirus Type 1; Rhinovirus Type 37; Feline calicivirus | 1 | Yes |
| Hydrogen peroxide | Accel TB Wipes | Virox Technologies Inc | Poliovirus | 1 | Yes |
| Hydrogen peroxide | Oxivir™ Wipes | Diversey Inc | Norovirus; Poliovirus Type 1; Rhinovirus Type 14 | 1 | Yes |
| Hydrogen peroxide | Oxivir 1 Wipes | Diversey Inc | Enterovirus Type D68 | 1 | Yes |
| Quaternary ammonium; Ethanol | Peak Disinfectant Wipes | North American Infection Control Ltd | Poliovirus Type 1; Rhinovirus | 1 | Yes |
| Sodium hypochlorite | Avert Sporicidal Disinfectant Cleaner Wipes | Diversey Inc | Human coronavirus | 1 | No |
| Quaternary ammonium; Isopropanol | Super Sani-Cloth Germicidal Disposable Wipe | Professional Disposables International Inc | Rhinovirus 39; Adenovirus 2 | 2 | Yes |
| Hydrogen peroxide | Clorox Commercial Solutions® Hydrogen Peroxide Cleaner Disinfectant Wipes | Clorox Professional Products Company | Norovirus | 2 | Yes |
| Quaternary ammonium | Nugen 2m Disinfectant Wipes | Lonza LLC | Human coronavirus | 2 | No |
| Quaternary ammonium | Caterpillar | Reckitt Benckiser | Human coronavirus | 2.5 | No |
| Quaternary ammonium; Isopropanol | Opti-Cide 3® Wipes | Micro-Scientific LLC | Rotavirus; Rhinovirus | 3 | Yes |
| Quaternary ammonium | AF3 Germicidal Disposable Wipe | Professional Disposables | Rotavirus; Adenovirus | 3 | Yes |

**Table S5, Part 2 of 2:** List of EPA-certified wipes. It is ordered by required contact time. All listed products are approved for healthcare use. If a product qualifies for the emerging viral pathogen claim, it is effective against a harder-to-kill virus than human coronavirus. All products on this list meet EPA’s criteria for use against SARS-CoV-2, including those marked as “No” in this column.

|  |  |  |  |  |  |
| --- | --- | --- | --- | --- | --- |
|  |  | International Inc |  |  |  |
| Quaternary ammonium; Ethanol; Isopropanol | Caviwipes 1 | Metrex Research | Adenovirus | 3 | Yes |
| Sodium hypochlorite | Caviwipes Bleach | Metrex Research | Feline calicivirus | 3 | Yes |
| Quaternary ammonium | PJW-622 | Clorox Professional Products Company | Rotavirus | 3 | Yes |
| Sodium hypochlorite | Clorox Healthcare® Bleach Germicidal Wipes | Clorox Professional Products Company | Canine Parvovirus; Feline Parvovirus | 3 | Yes |
| Quaternary ammonium; Ethanol; Isopropanol | Sani-Cloth Prime Germicidal Disposable Wipe | Professional Disposables International Inc | Feline Calicivirus | 3 | Yes |
| Quaternary ammonium | Sani-cloth Germicidal Disposable Cloth | Professional Disposables International Inc | Human coronavirus | 3 | No |
| Quaternary ammonium | Clorox Disinfecting Wipes | The Clorox Company | Rotavirus | 4 | Yes |
| Quaternary ammonium | Clorox Commercial Solutions® Clorox® Disinfecting Wipes | Clorox Professional Products Company | Rotavirus | 4 | Yes |
| Quaternary ammonium | Lonza Disinfectant Wipes Plus | Lonza LLC | Human coronavirus | 4 | No |
| Quaternary ammonium | Nugen Low Streak Disinfectant Wipes | Lonza LLC | Human coronavirus | 4 | No |
| Quaternary ammonium | Clorox Healthcare® VersaSure® Wipes | Clorox Professional Products Company | Norovirus | 5 | Yes |
| Ethanol | PURELL Professional Surface Disinfectant Wipes | GOJO Industries Inc | Norovirus | 5 | Yes |
| Phenolic | Sporicidin (Brand) Disinfectant Towelettes | Contec Inc | Human coronavirus | 5 | No |

## 14.2 Appendix B Suggested Decontamination Protocols

1. Perform hand hygiene.
2. Put on a gown, gloves, and a protective mask.
3. Wipe the surface of the filter with 70% ethanol or hydrogen peroxide, and carefully remove the filter.
4. Discard the wipe as biohazardous waste. Discard the filter if it is intended for single-use only. If you plan to reuse the filter, follow the instructions from the manufacturer of the filter to store it properly, and DO NOT WET the surface of the filter.
5. Wash mask and filter adapter thoroughly in warm water (43 deg C [110 deg F] maximum) with a mild detergent. A stiff bristle (not wire) brush may be used to facilitate the removal of dirt. Make sure to clean all the valves to avoid potential clogged valves.
6. Rinse the mask and adapter thoroughly in clean, warm (43 deg C [110 deg F] maximum), running water. Drain.
7. Prepare a 50ppm chlorine solution: add one milliliter of laundry bleach per liter of warm water (43 deg C [110 deg F] maximum). Prepare enough solution to fully immerse the entire mask. Remove all the air in the side channel to ensure full immersion. Please note that mixing bleach solutions with detergents can generate toxic substances. Make sure to wash out all the detergents before immersing the mask and adapter into bleach.
8. Immerse the mask and adapter in the hypochlorite solution (50 ppm of chlorine) for 2 minutes.
9. Wearing fresh gloves, rinse the mask and adapter thoroughly in clean, warm, running water (43 deg C [110 deg F] maximum). The mask and adapter must be thoroughly rinsed with water to remove any detergents or disinfectants that may result in dermatitis or damage of the mask components.
10. Dry the mask and adapter with a clean lint-free cloth or allow to air dry.
11. Reassemble the mask and store in a clean space.

*This protocol is based on OSHA’s procedure for cleaning respirators:* https://www.osha.gov/laws-regs/regulations/standardnumber/1910/1910.134AppB2

## 14.3 Appendix C Communication App User Instructions

### 14.3.1 Current Android app download instructions

1. Download the beta version of the app by visiting http://kylecombes.com/app-debug.apk on your phone.
2. Open the download. Your phone should prompt you to allow installations from unknown (i.e. non-Google Play) sources. If it does, simply follow the on-screen instructions. If no such prompt appears and the installation merely fails, do the following:
  - Allow app installations from unknown sources by going to your phone’s Settings -> Security -> Unknown Sources and enable installations. This setting might have a different name, depending on your version of Android and device manufacturer.
  - Try to install the app again by re-opening the download or revisiting the link above.

#### 14.3.2 App usage instructions

1. Connect your phone to a Bluetooth device with a microphone. We recommend using cheap Bluetooth headphones, which can be put inside a sterilized plastic bag, and placed inside the face compartment of the snorkel mask.
2. (Optional, for extra amplification) Connect an external speaker to the phone’s wired headphone jack using an aux cord. If you do not do this, the phone will use its internal speakers.
3. Open the app.
4. If the app detects a Bluetooth device capable of streaming audio, it will say “Bluetooth ready.” If it does not, follow the on-screen instructions to ensure your device is connected properly.
5. Click “Start” to begin relaying audio from the microphone to the speaker.
6. You may now turn off the phone’s screen or switch to another app if you would like. If you need to switch speakers at any point, click “Stop,” do any plugging/unplugging, and then click “Start.”
7. Click “Stop” end relaying audio.

##### 14.3.3 Troubleshooting

If the sound is playing out of the Bluetooth speaker instead of through the wired speaker, simply unplug and replug in the wired speaker.

Currently, the iOS version of this app is under development, and the Android app is pending publication in the Google Play Store. Please reach out to the provided contacts if you have any feedback, questions, or concerns. Contacts: Emma Pan (epan549 at gmail.com), Kyle Combes (kcombes at olin.edu)

## 14.4 Appendix D

Suggested Donning and Doffing procedures

FURTHER REVIEW OF THIS PROTOCOL BY INFECTION DISEASE SPECIALISTS OR EHS OFFICERS IN HOSPITALS IS REQUIRED.

### 14.4.1 Donning Procedures

1. Eat, drink and go to the restroom. Check everything that you need to bring into the patient’s room is available.
2. Tie up your hair and remove unnecessary accessories on wrists, neck and ears.
3. Perform hand hygiene.
4. Put on a surgical cap, tucking loose hair away from the face and neck.
5. Put on the first layer of gloves.
6. Put on your gown by placing your head through the top of the opening and insert your arms into the sleeves. Tie the gown properly.
7. Carefully inspect the disinfected snorkel mask, adapter and viral filter of Pneumask.
8. (Optional if they are connected already) Serially connect the adapter and viral filter to the snorkel mask.
9. (Optional for Bluetooth user) Turn on the Bluetooth microphone. Check the connection with the smartphone. Put the Bluetooth microphone in a clean, small ziplock bag, Tape and secure the ziplock bag inside the mouth chamber without blocking the chin valve.
10. Perform a suction test: put the mask to your face, press it slightly while inhaling to create a suction between the mask and your face. The mask should suction to your face. Try smiling and see if the movement of facial muscle breaks the seal. IF MASK DOES NOT PASS SUCTION TEST, THE MASK SHOULD NOT BE USED.
11. Put on the mask fully. Adjust and tighten the straps as needed.
12. Inhaling or exhaling while closing off the top of the filter with your hand can be done for another seal check. With a good facial seal, the mask should suck down on your face with a good seal with inspiration, and not lift off face with exhalation.
13. Perform hand hygiene.
14. Place your gloved thumbs through the loops of the gown. Put on the second layer of gloves.

#### 14.3.2 Doffing Procedures

1. In the anteroom (or the doorway if there is no anteroom), grasp the gown and pull away from your body. Carefully fold the gown into ball-shape and only touch the outer surface of the gown with gloved hands. Remove the gown and the outer gloves together. Dispose the gown and the gloves into a dedicated trash can.
2. Perform hand hygiene (you should still be wearing your inner layer of gloves).
3. Leave the anteroom.
4. Perform hand hygiene.
5. Inspect the integrity of the inner glove. If the inner glove is intact without gross contamination, proceed through the next step. If the inner glove is torn, broken, or grossly contaminated, remove the inner glove. Perform hand hygiene, and put on a new pair of gloves.
6. Lean head forward into sniff position. Loosen the straps and remove the mask without touching your face.
7. Use EPA-approved alcohol-based wipe, bleach wipe or hydrogen peroxide wipe to wipe the outer surface of the mask, adapter and filter. Dispose the wipe. The required contact time for different EPA-approved wipes can be found in Appendix A Table S5.
8. Use EPA-approved alcohol-based wipe, bleach wipe or hydrogen peroxide wipe to wipe the inner surface edges of the mask. Dispose the wipe.
9. Gently disconnect the filter from the mask. Dispose the filter if it is designed for one-time usage. If you plan to reuse the filter, follow the instructions from the manufacturer of the filter to store it properly, and DO NOT WET the surface of the filter.
10. Put the mask into a clean dedicated box.
11. Perform hand hygiene.
12. Remove the surgical cap.
13. Perform hand hygiene.
14. Remove the inner gloves.
15. Perform hand hygiene.
16. (Optional for Bluetooth user) Remove the ziplock bag which contains the Bluetooth microphone. Wipe the surface of the ziplock bag with EPA-approved wipe (Appendix A Table 5) and open the ziplock bag. Perform hand hygiene. Dump the microphone from the ziplock bag with one hand (dirty hand) onto the other hand (clean hand). Dispose the ziplock bag with your dirty hand. Place the microphone somewhere you will not forget with your clean hand. Perform hand hygiene.
17. By the end of the shift, bring the box and the mask to somewhere you can fully decontaminate it following our decontamination protocol, or hand it to hospital technicians per hospital policy. The recommendation of using disinfection wipe between patients and fully washing it after one shift is following the recommendation of NIOSH on elastomeric respirator.

## 14.5 Appendix E

Supplementary Figures

**Figure S1a.**
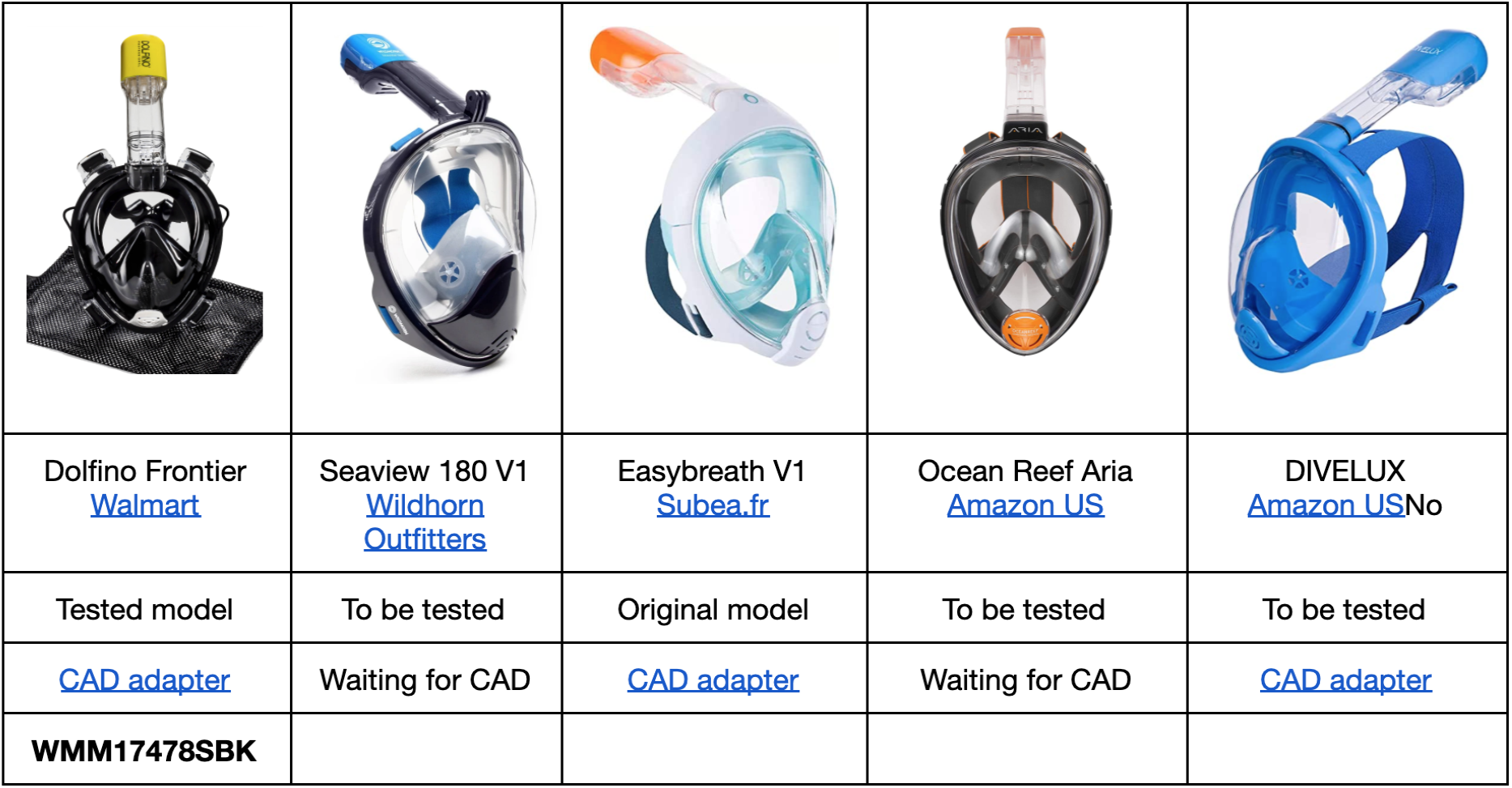
Table of various full-face snorkel masks readily available in the market.

**Figure 28.**
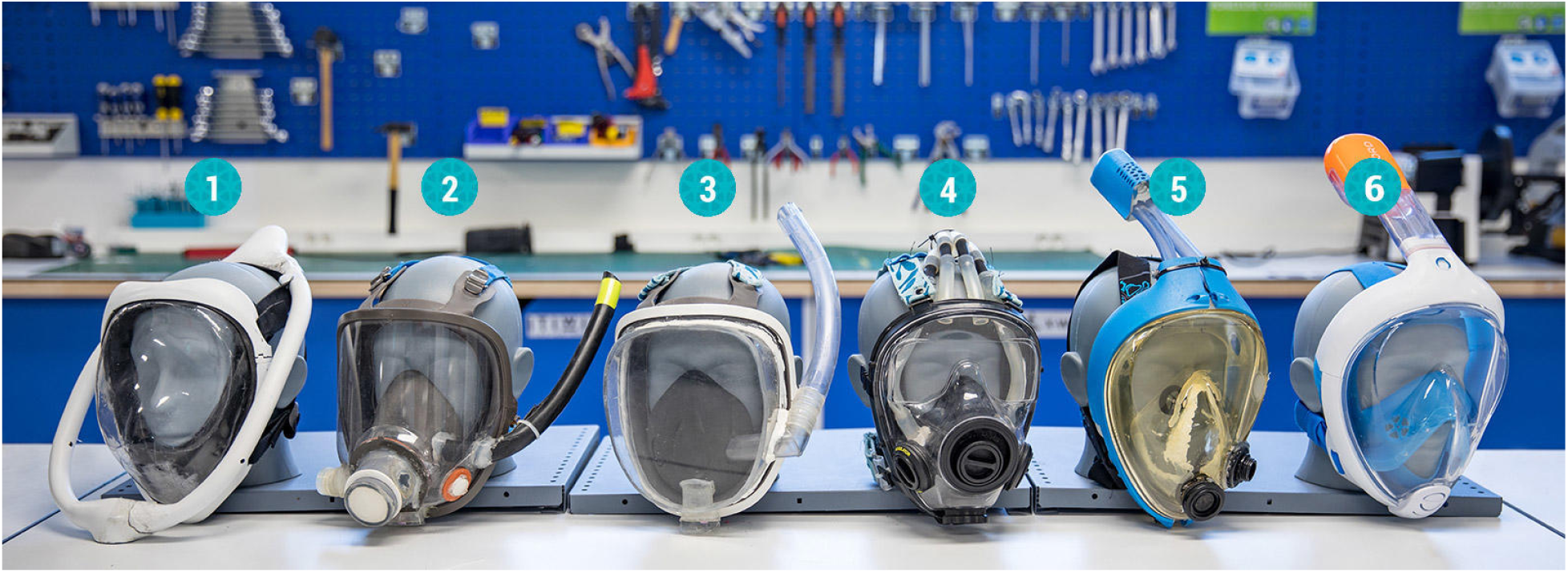
**Figure S1b** Evolution of the prototype of full-face snorkel masks over time by the original inventors - Subea Team.

**Figure 29.**
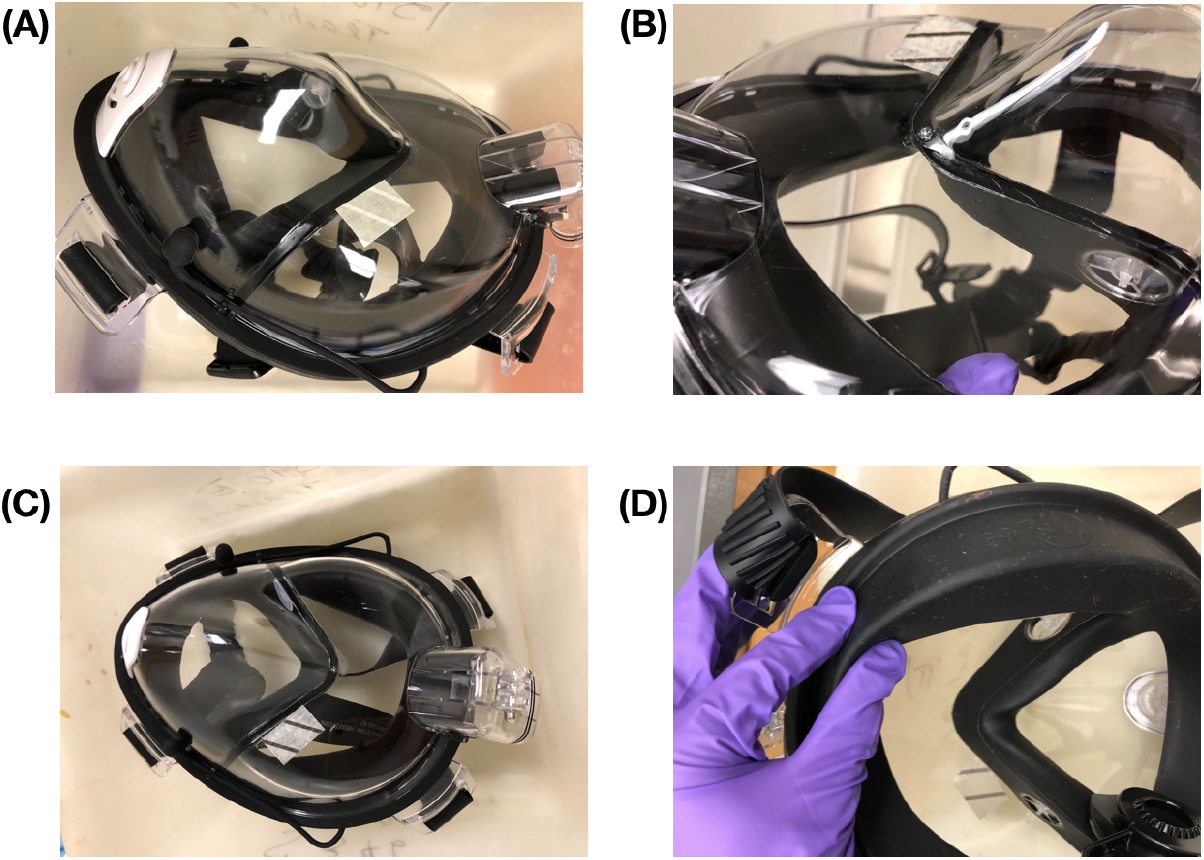
**Figure S2** Before Autoclave (A), Round 1 (B), Round 2 (C) Round 3 photos and [video](https://drive.google.com/file/d/16njaEgjBuuFCA5-InksTqqGLpsnHS2kR/view) (D). The mask survives autoclaving at 121 deg C and 15psi for a total of 90 minute cycle (done in three cycles of hot and room temperature) without apparent damage. The folders of photos and/or videos are available with the link in the caption.

**Figure 30.**
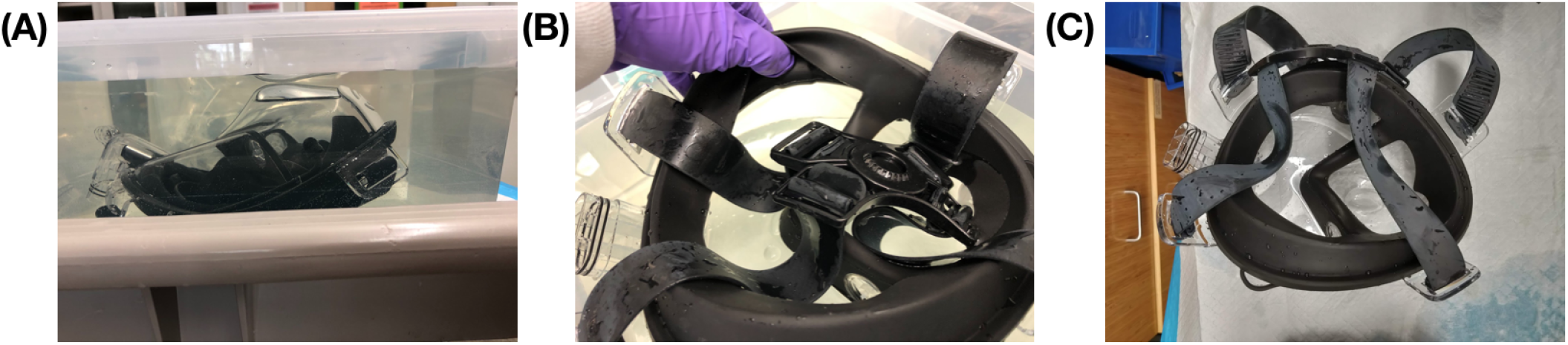
**Figure S3**: (A) Mask immersed in 10% bleach in a simple tupperware. (B) After 2 hours of immersion, we found no apparent damage to the mask. (C) After 8.25 additional hours of immersion, we again observed no apparent damage to the mask, though white coating had formed on the surface of the black elastomer bands.

**Figure 31.**
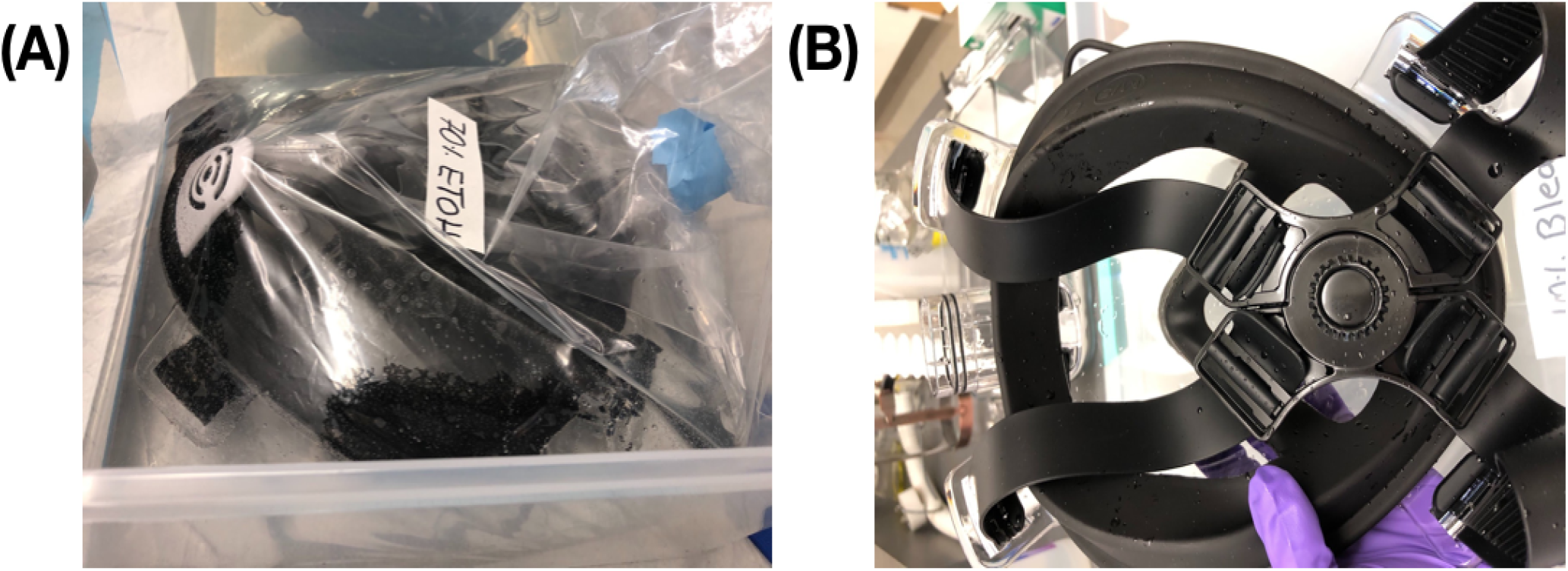
**Figure S4**: (A) Before (B) After 2 hours of immersion, we found no apparent functional damage to the mask.

**Figure 32.**
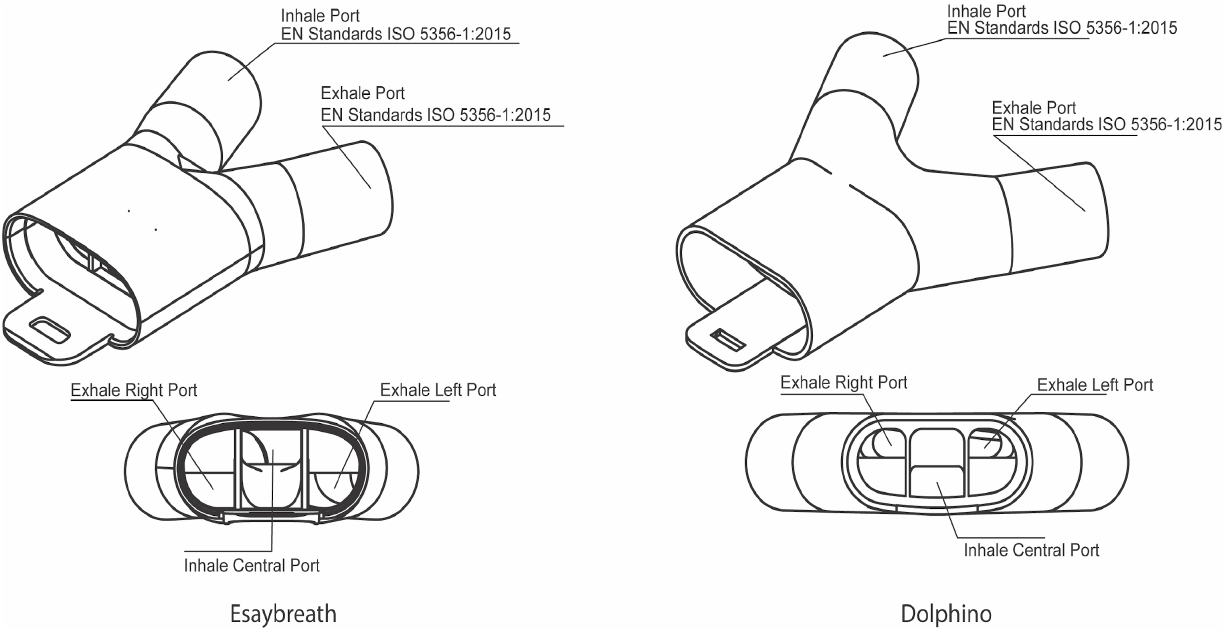
**Figure S5**:Dolphino and Easybreath double port inhale/exhale separation adapters. The design followed EN Standards ISO 5356-1:2015. A set of connectors ISO 5356 are available on GrabCAD thanks to Filip Kober. The CAD files of the universal adapter for Easybreath (https://a360.co/2Ukh6w5) and Dolphino Frontier (https://a360.co/39jnJCQ) are available with the links provided.

**Figure 33.**
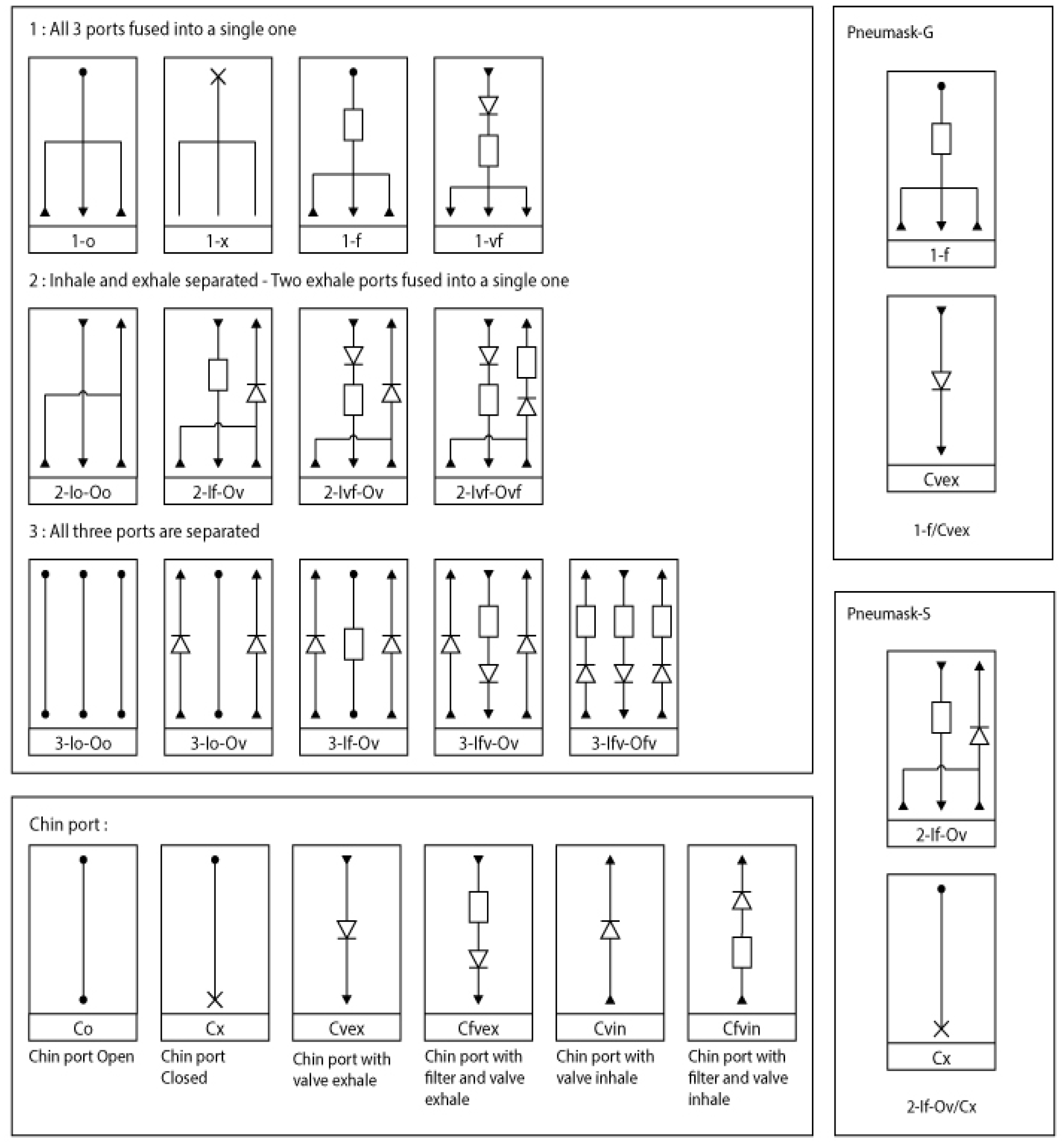
**Figure S6**: We have developed a nomenclature of the different combinations existing for the air flow pathway. Pneumask-G could then be named 1-f/Cvex and Pneumask-S named 2-If-Ov/Cx. Notation: diodes = one-way valves; resistor = filter. Please find the .ai original vector file [here](https://drive.google.com/file/d/14oqTFKCKgeTwWTWVno7Zt8eip-uasduT/view).

